# Preventable deaths involving medicines in England and Wales, 2013-22: a systematic case series of coroners’ reports

**DOI:** 10.1101/2022.11.01.22281803

**Authors:** Harrison S France, Jeffrey K Aronson, Carl Heneghan, Robin E Ferner, Anthony R Cox, Georgia C Richards

## Abstract

**Objectives:** To identify medicines-related deaths in coroners’ reports and to explore concerns to prevent future deaths.

**Design:** Retrospective case series of coroners’ Prevention of Future Deaths reports (PFDs).

**Setting:** England and Wales.

**Participants:** Individuals identified in 3897 PFDs dated between 1 July 2013 and 23 February 2022, collected from the UK’s Courts and Tribunals Judiciary website using web scraping, and populated into an openly available database: https://preventabledeathstracker.net/

**Main outcome measures:** Proportion of PFDs in which coroners reported that a therapeutic medicine or drug of abuse caused or contributed to a death; characteristics of the included PFDs; coroners’ concerns; recipients of PFDs and the timeliness of their responses.

**Results:** 704 PFDs (18%; 716 deaths) involved medicines, representing an estimated 19,740 years of life lost (average of 50 years lost per death). Opioids (22%), antidepressants (9.7%), and hypnotics (9.2%) were the most common drugs involved. Coroners expressed 1249 concerns, primarily related to patient safety (29%) and communication (26%), including failures of monitoring (10%) and poor communication between organisations (7.5%). NHS England (6%), the Department of Health and Social Care (5%) and the Medicines and Healthcare products Regulatory Agency (2%) received the most medicines-related PFDs. However, most expected responses to PFDs (51%; 615/1245) were not reported on the UK’s Courts and Tribunals Judiciary website.

**Conclusions:** One in five deaths deemed preventable by coroners involved medicines. Taking actions to address coroners’ concerns, including improving patient safety and poor communication, should increase the safety of medicines. Many concerns were raised repeatedly, but half of PFD recipients failed to respond, suggesting that lessons are not generally learned. The rich information in PFDs should be used to foster a learning environment in clinical practice that may help reduce preventable deaths.

**Trial registration:** https://doi.org/10.17605/OSF.IO/TX3CS

**Summary box:** *What is already known on this topic?:* Medicines are essential to the provision of healthcare, but if used inappropriately, have the potential to cause significant harms, including death. When an unnatural death occurs, these deaths are often reported to the coroner, which can result in a report to highlight concerns to prevent future deaths. Samples of coroners’ reports have been analysed to identify concerns relating to preventable deaths involving medicines, which found that anticoagulants contributed the most. However, an investigation of all available reports has not been conducted to determine the overall impact of medicines.

*What this study adds?:* One in five preventable deaths in England and Wales involved a medicine or drug of misuse, costing nearly 20,000 years of life lost. Opioids, antidepressants, and hypnotics were the most common medicines involved in preventable deaths. Coroners repeatedly raised similar concerns, primarily relating to patient safety and communication. However, it is unclear whether these reports are being used in clinical practice to guide actions to prevent similar deaths.

## Introduction

One in four (26%) of the UK population receives a prescription for at least one medicine yearly (1). The aging population and corresponding multimorbidity (2) imply that the use of medicines is expected to increase further. In 2013, drugs introduced since 1981 were estimated to have saved 149 million years of life in 22 countries (3). However, drugs are also associated with harms, which can result from medication errors, adverse drug reactions, and misuse. They have been implicated in 7-18% of hospital deaths in Spanish studies (4,5), and avoidable adverse drug events contribute to over 1708 deaths annually in England (6). Investigating and understanding the issues underlying medicine-related deaths may provide useful information for practitioners and policymakers and help improve patient safety.

Since 1984, coroners in England and Wales have had a duty to report details of deaths when the coroner believes that actions should be taken to prevent future deaths (7–9). These reports, named Prevention of Future Deaths reports (PFDs), are mandated under UK law, and require recipients to respond within 56 days. A study that examined a sample of PFDs between 24 April 2015 and 7 September 2016 showed that medicines contributed to 20% of deaths (99 PFDs of 500 included) deemed preventable by coroners in England and Wales (10,11). Other studies of PFDs have provided insights into factors related to deaths related to specific medicine types, including anticoagulants and medicines purchased online (12,13). However, an analysis of all available PFDs has not been conducted to determine the overall contributions of medicines to preventable deaths. Therefore, we aimed to conduct a systematic case series of all available PFDs, to identify and characterise deaths involving medicines and to explore concerns to prevent future deaths.

## Materials and Methods

### Study design and data sources

We designed a retrospective case series and pre-registered the study protocol on the Open Science Framework (OSF) (14). Data were acquired from the Courts and Tribunals Judiciary website (15) using web scraping to populate a table for manual screening, as described elsewhere (12,16). The web scraper produced a database, called the Preventable Deaths Database (https://preventabledeathstracker.net/database/), that included 3897 PFDs as of 23 February 2022.

### Data screening and eligibility

We screened the 3897 PFDs using a pre-defined definition of medicines-related deaths developed from previous work (see Supplement 1) and included PFDs if a medicine caused or contributed to death. We excluded PFDs in which the sole agent was alcohol or tobacco, and cases in which a medicine or a component of the therapeutic process were not deemed to have contributed to or caused death, or when only delays in assessment, investigation, or administration contributed to a death.

### Data extraction

After screening, data relating to the characteristics of the deceased, circumstances of the death, coroners’ concerns, and responses from recipients were extracted from included PFDs (see Table S1 in Supplement 1 for a full list of fields). The web scraper automatically extracted some information from PFDs and presented it in the Preventable Deaths Tracker (17). The types of medicines were classified by their generic names under BNF guidance (18). If a medicine was missing from the BNF, classification was provided by the International Union of Basic and Clinical Pharmacology/British Pharmacological Society (IUPHAR/BPS) Guide to Pharmacology (19) or, failing that, the Misuse of Drugs Act 1971 (20).

### Data Analysis

The number of included PFDs and their frequencies as a proportion of all PFDs were plotted over time. Frequencies were reported for categorical variables (sex, coroner’s area, type of medicine, source of medicine) and medians and interquartile ranges (IQR) were calculated for continuous variables (age, latency from death, number of medicines contributing to death). We analysed the sex distribution with a one-tailed binomial test. We estimated years of life lost (YLL) using the formula *YLL = EC* − ∑ *A*; where *E* = life expectancy (years; set using the Office of National Statistics (ONS) average life expectancy for males (79 years) and females (82.9 years (21)); *C* = the number of subjects with age below the life expectancy; and *A* = age of the subject at death, adapted from the World Health Organization’s (WHO’s) formula (22), for males and females independently and combined.

To identify geographical trends in reporting, we ranked coroners’ areas by deciles, based on the raw number of medicines-related PFDs produced and the proportion of all PFDs from each area that were medicines-related, to reduce the influence of differences in coroners’ reporting trends.

We used directed content analysis to collate and evaluate coroners’ concerns (23). We used a framework of concerns developed from previous publications (10,12,24) to categorise concerns into major and minor themes. If concerns failed to fit into predefined categories, we developed new themes.

To calculate response rates, we used the 56-day legal requirement to classify responses as “early” (>7 days before the due date), “on-time” (≤7 days before or after the due date), “late” (>7 days after the due date), or “overdue” (response was not available on the Judiciary website as of 23 February). The response rates were grouped by the classes of individuals and organizations to whom PFDs were addressed. Recipient classes were determined through conventional content analysis, identifying common groups.

### Software and data sharing

The study protocol was preregistered on the OSF (14) and all study materials, data, and code are openly available via GitHub (25) and the OSF (26). We used SPSS (version 28) to conduct statistical analyses and Data Wrapper© to create all the figures.

### Protocol deviations

We did not report analyses on the duration of the inquest, setting of death, or “action should be taken” recommendations from the PFDs, as the information was often incomplete. We also increased the study’s duration and data availability, from 28 June 2021 to 23 February 2022.

### Patient and public involvement

No patients were directly involved in this study. However, the public is continuously engaging and involved with the broader programme of research via the Preventable Deaths Tracker website: https://preventabledeathstracker.net/. The lead author (HSF) presented preliminary findings at the UK Clinical Pharmacology Colloquium (November 2021) and during World Smart Medication Day 2022 for the British Pharmacological Society and the International Union of Basic and Clinical Pharmacology poster competition (May 2022), which was subsequently published in *Pharmacology Matters* (27), and authors (HSF and GCR) presented some key findings at the EBM Live 2022 Conference for further international engagement (July 2022).

## Results

There were 704 medicines-related PFDs eligible for inclusion (18% of all PFDs), representing 716 individual deaths (Fig. 1). The reporting of medicines-related PFDs increased by 8% from 15% in 2013 to 23% in 2022 (Fig. 2; Table S2 in Supplement).

**Figure 1:**
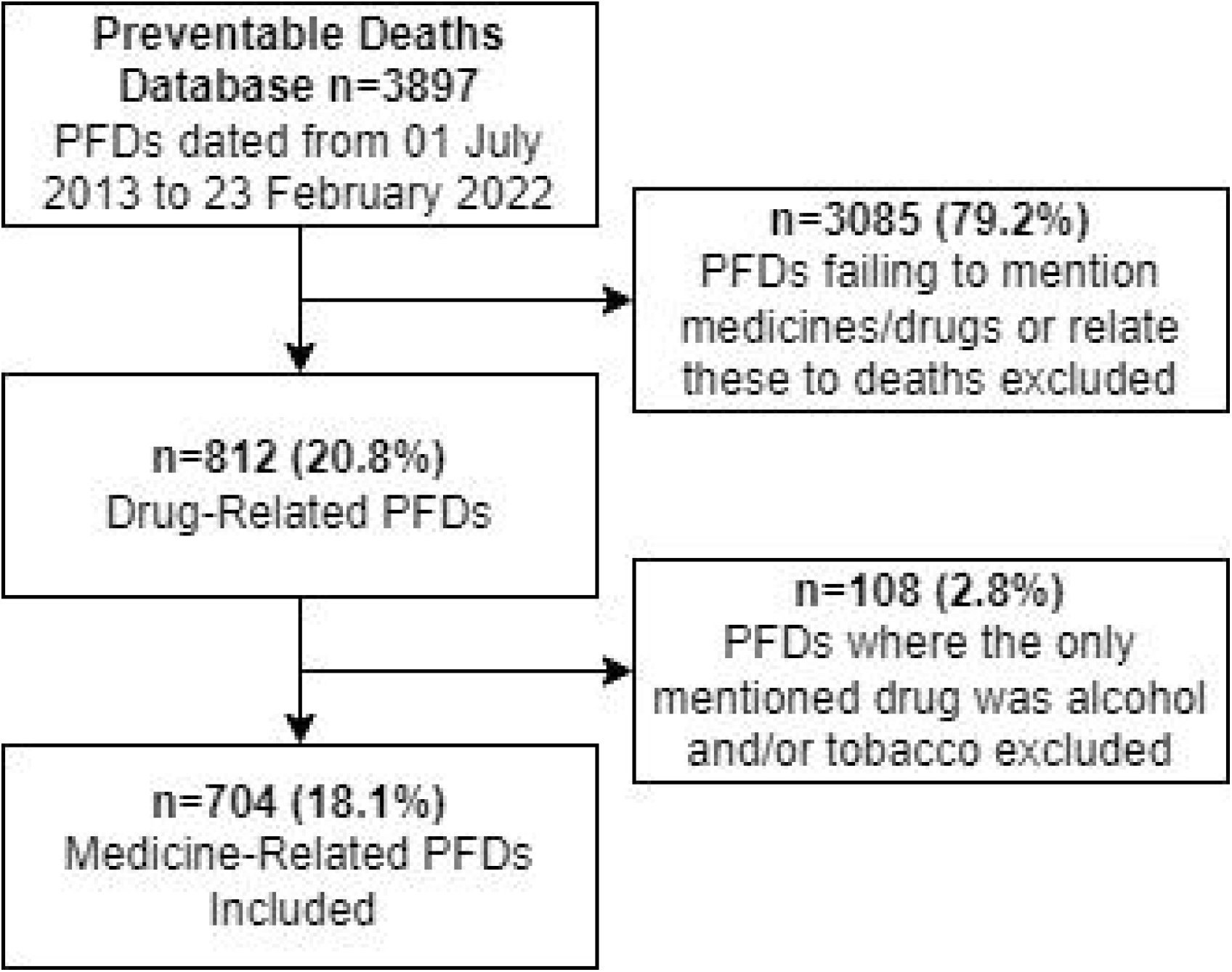
Flow diagram of the screening of Prevention of Future Deaths reports (PFDs) in England and Wales from the Preventable Deaths Database using the eligibility criteria for medicine-related deaths.

**Figure 2:**
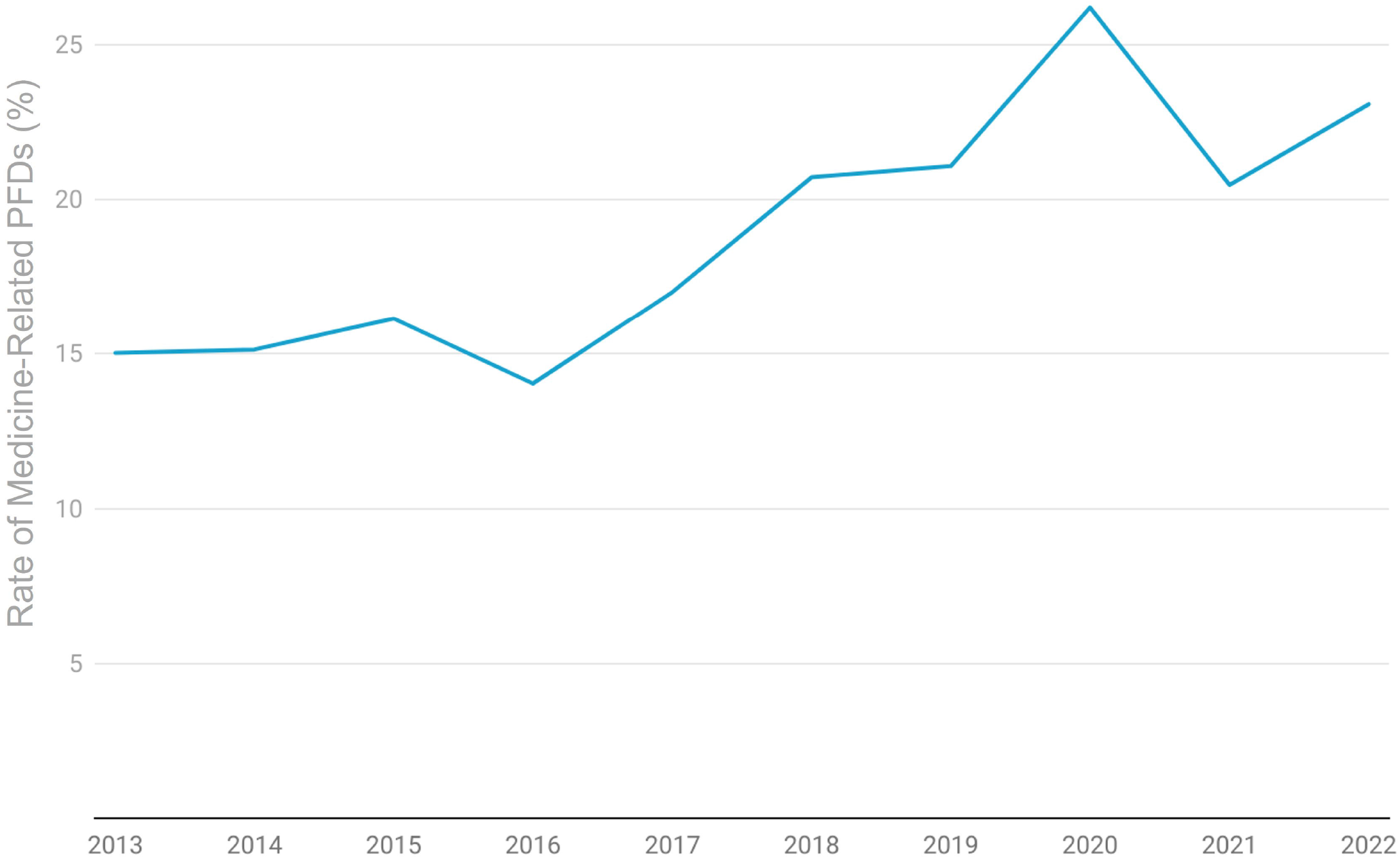
Rate of medicine-related Prevention of Future Deaths reports (PFDs) in England and Wales between July 2013 and 23 February 2022, calculated as the percentage of reports meeting the inclusion criteria divided by the total number of PFDs published each year.

Of the 396 deaths that reported ages below the life expectancy, 19 740 estimated years of life were lost, equating to an average of 50 years of life lost for every medicine-related preventable death (median age: 42 years, IQR: 29-63 years, n=450). Most deaths (68%, n=480) involved males (P<0.01) and reporting varied geographically.

On the Judiciary website, PFDs are classified into report types based on the causes of death. For all medicines-related PFDs, only 28% (n=197) were classified as “Alcohol, drug, and medication related deaths” and 3% (n=22) were “Product related deaths”.

In 45% (n=319) of medicine-related PFDs, coroners reported medical histories and information on social histories; in 40% of PFDs (n=280) coroners reported mental health problems (Fig. S1 and Table S3 A-C in Supplement 1).

Coroners in Manchester, Birmingham, and Inner London wrote the most medicines-related PFDs (Fig 3; Table S4 in Supplement). No PFDs were reported in 13 coroner areas: Carmarthenshire and Pembroke; Ceredigion; North Lincolnshire and Grimsby; North Northumberland; North Tyneside; Northwest Kent; Northwest Wales; North Yorkshire (Eastern and Western); Rutland and North Leicestershire; Sefton, Knowsley, St Helens; South Northumberland; and Sunderland.

**Figure 3:**
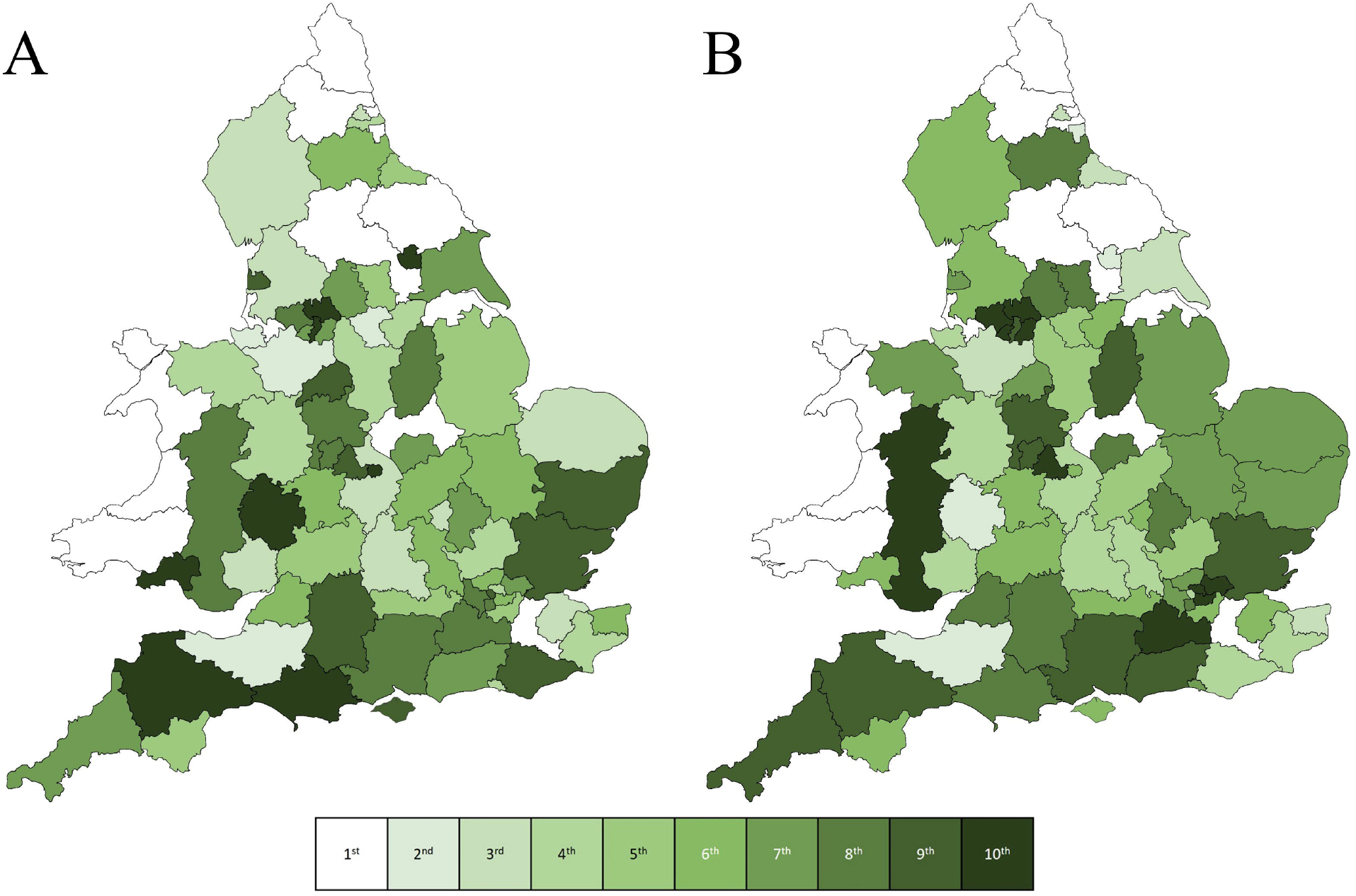
Map of coroners’ areas in England and Wales displaying the distributions of the 704 medicines-related Prevention of Future Death reports (PFDs) published between July 2013 and 23 February 2022 by raw number **(A)** and rates of medicine-related PFDs by all PFDs in each area **(B)**. Each area was graded into deciles, from no PFDs (white) to highest (dark green).

### Types of medicines

1172 medicines were reported in the 704 PFDs; however, most (65%) reported one medicine (median: 1; IQR:1-2, range: 1-12; Table S5 in Supplement). Opioids (22%), antidepressants (9.7%), and hypnotics (9.2%) were the most common drugs involved in deaths (Fig. 4; Table S6 in Supplement 1).

**Figure 4:**
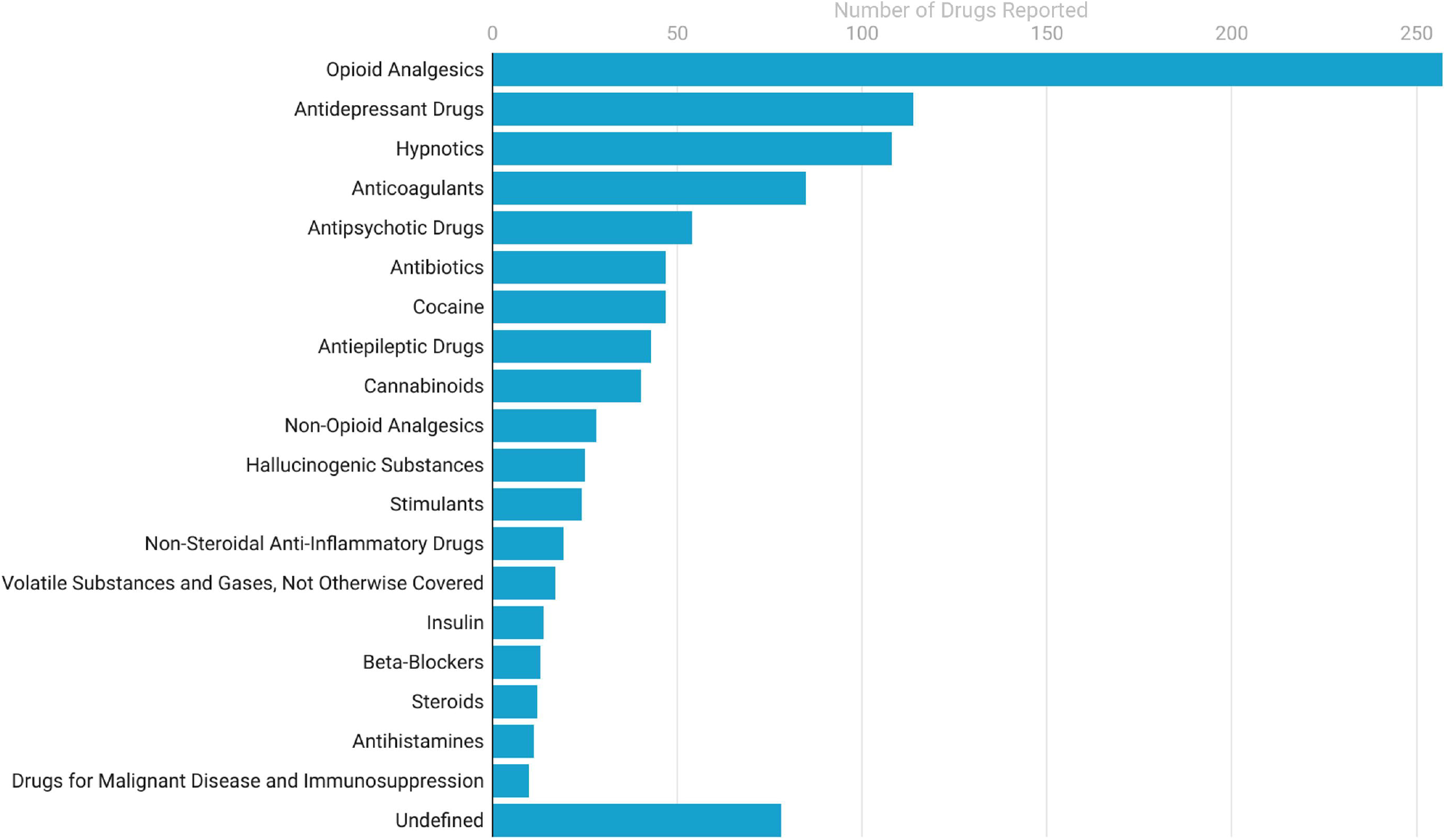
Frequencies of the 20 most common types of medicines from 704 included medicines-related Prevention of Future Death reports (PFDs) published between July 2013 and 23 February 2022; Coroners reported ‘undefined’ medicines without name or class provided.

In 93% (n=652) of PFDs coroners reported the source(s) of medicine(s). Most (69%, n=450) were prescribed, and other common sources were illicitly acquired medicines (31%, n=201), over-the-counter medicines (5.7%, n=37), ‘legal highs’ (2.3%, n=15), and medicines prescribed for someone other than the deceased (1.7%, n=11). 32 PFDs (5.6%) involved substances obtained using the internet.

### Coroners’ concerns

We identified 1249 concerns reported by coroners. We categorised the concerns into six major themes (Patient Safety; Communication; Education & Training; Resources; Regulations; and Failures to Carry Out Necessary Tasks/Protocols/Jobs) plus 44 minor themes. Failure to monitor/observe patients was the most common concern (10%, n=133), followed by poor communication between organizations (7.5%; n=99), unsafe protocols (7.3% n=96), and failure to keep accurate medical records/care plans (6.9%, n=90) (Fig. 5). Illustrative examples of deaths and associated coroners’ concerns are provided in Supplement 2.

**Figure 5:**
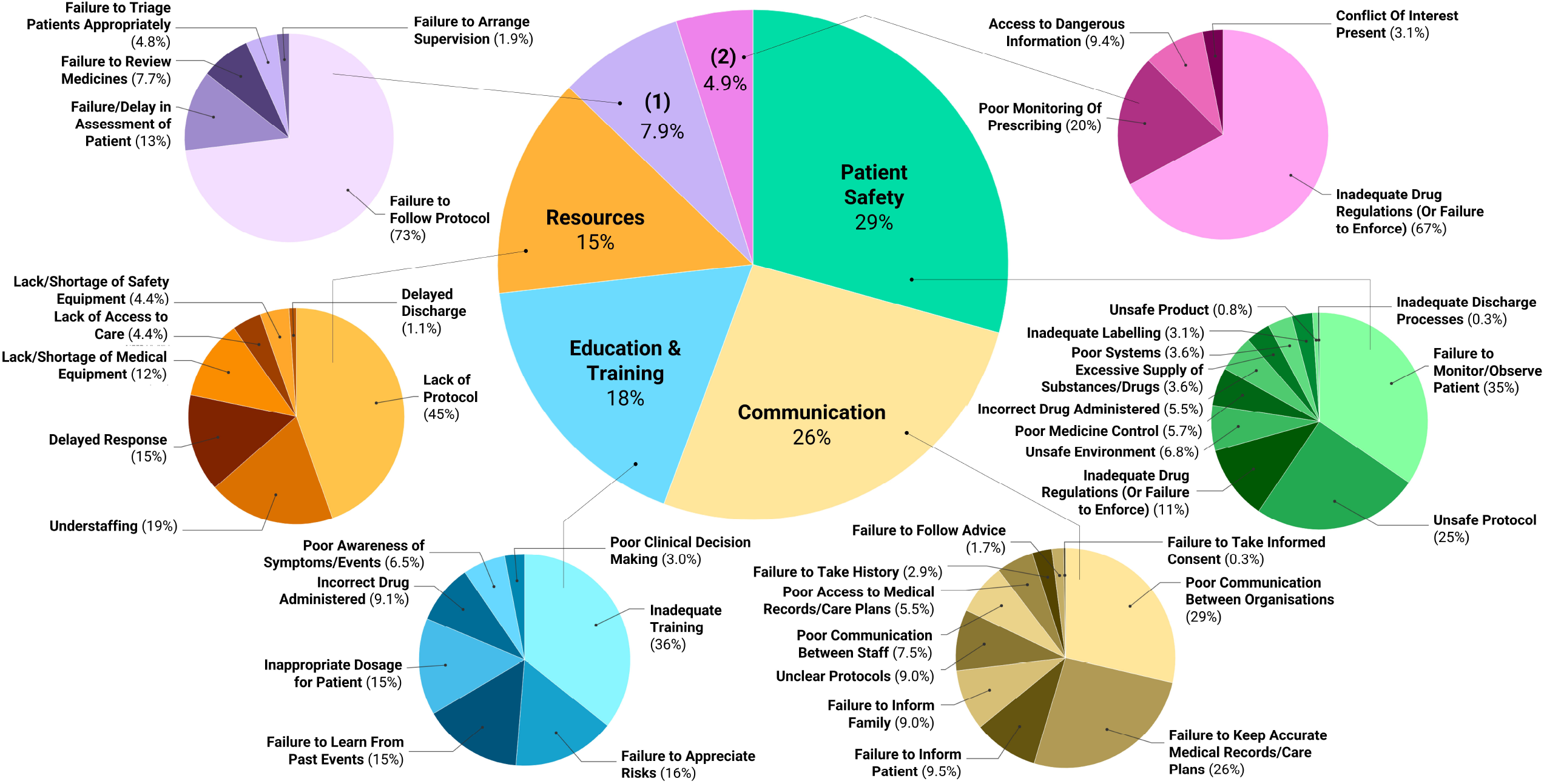
Coroners’ concerns from 704 included medicines-related Prevention of Future Death reports (PFDs) in England and Wales published between July 2013 and 23 February 2022, grouped into major and minor themes using directed content analysis. The purple sixth (1) represents the failure to carry out necessary tasks/protocols/jobs and the pink sixth (2) represents regulation.

### Responses to PFDs

Coroners sent the 704 medicine-related PFDs to 1245 individuals and organizations, representing 564 unique recipients. The most common recipients were NHS England (6% of all reports; n=77), followed by the Department of Health and Social Care (DHSC; 5%; n=67) and the Medicines and Healthcare products Regulatory Agency (MHRA; 2%; n=26) (Fig. S2 in Supplement 1). Only 49% (n= 615) of recipients responded; of those, 25% (n=152) were early, 52% (n=314) on time, and 24% (n=149) late (Fig. 6; Table S7 in Supplement 1). At the time of analysis, 630 responses (51%) remained overdue across 268 individual PFDs.

**Figure 6:**
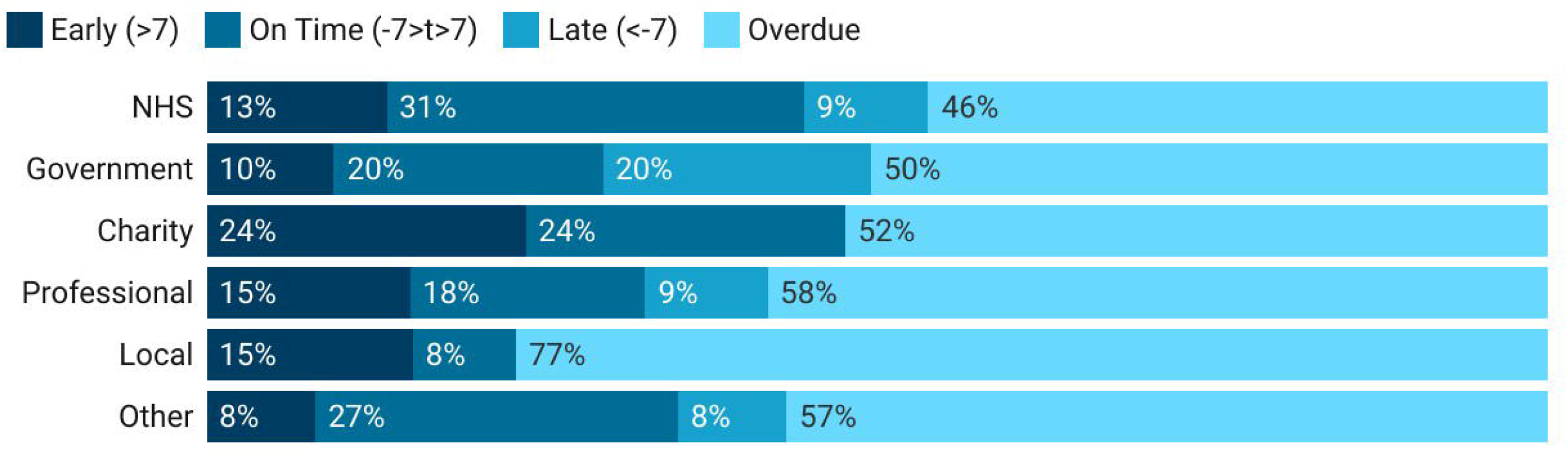
Proportions of responses to medicines-related Prevention of Future Death reports (PFDs) in England and Wales between July 2013 and 23 February 2022, using the 56-day legal requirement to classify responses as “early” (>7 days before due date), “on-time” (≤7 days before or after the due date), “late” (>7 days after the due date), or “overdue” (response was not available on the Judiciary website as of 23 February) and reporting results by organization type.

## Discussion

There were 716 deaths involving medicines deemed preventable by coroners in England and Wales reported between July 2013 and 23 February 2022, amounting to nearly 20,000 estimated years of life lost. One in five medicines-related PFDs involved opioids and one in ten involved antidepressants. Coroners in Manchester and Inner London wrote the most medicines-related PFDs, whereas many in Northeast England and West Wales wrote no PFDs. Coroners raised repeated concerns, often relating to patient safety and communication. Despite statutory obligations for recipients to respond within 56 days, half of the legally required responses were overdue at the time of analysis.

### Strengths and limitations of the study

Our study built on previous work (10), using a reproducible method to collect all available PFDs (16). Nevertheless, this PFD population cannot represent all preventable deaths in England and Wales, since 153,008 deaths were deemed avoidable across Great Britain in 2020 (28) and 40% of UK deaths are reported to coroners, 14% progressing to inquest (29). However, our findings identified repeated concerns that have national importance for preventing future deaths. Although not all lessons can be generalisable internationally, similar systems exist in New Zealand (30), Australia (31,32), and Canada (33). Thus, our reproducible methods have applications across continents.

There are no quality control mechanisms to examine the content of PFDs, which creates heterogeneity and limits the application of our findings. We identified missing information across most variables, including age (missing in 36% of PFDs), types of medicines (missing in 7%), and dates of responses. Some missing information may depend on coroners’ reporting practices, which we controlled for where possible. For example, geographical variation changed after controlling for the total number of PFDs by area. We also found inconsistencies on the Judiciary website. For example, only 28% of medicine-related PFDs were categorised as “Alcohol, drug and medication related deaths”, which increases the time needed to manually screen PFDs for inclusion. The lack of guidance on when a PFD should be written may underlie these problems. Thus, guidelines may help ensure the utility of PFDs in preventing future deaths.

### Comparison with previous literature

Compared with previous investigations of PFDs that found anticoagulants as the most common drugs involved in deaths (10,12), we found that opioids were dominant. This difference can be attributed to the quality of our data collection methods, which allowed us to examine all available PFDs to which medicines contributed. Compared with previous studies (10–12,24,34), concerns reported by coroners were the same, except for those relating to patient safety, which has not been identified in PFDs before. However, in a study examining hospital incident reports, 18% of concerns were related to patient safety (35). As in previous case series of PFDs, we found a dominance in male-related deaths (10,34,36,37) and identified geographical variations in writing and responding to PFDs (12,34,37,38). Although responses to PFDs are required by law, research continually highlights low response rates and the need for reform (11,12,37,39), so that PFDs can stimulate significant widespread healthcare changes.

### Interpretation and implications for clinicians and policymakers

Our research identifies over 1200 concerns reported by coroners that if not addressed will cause further harms and repeat preventable deaths. The concerns we have identified provide lessons for clinical practice, including the need for better monitoring and observation of at-risk patients in hospitals as well as regular medication reviews in the community. Extra considerations are needed to reduce omissions in record keeping and miscommunications between organisations, particularly when discharging or reviewing patients receiving medications. We also discovered a need for enhanced training and education for those caring for patients taking medicines, especially when controlled substances such as opioids are involved. Coroners also acknowledged a lack of resources that led to understaffing, equipment requirements, and need for policies and guidelines to help with decision making in complex scenarios involving medicines. However, our study did not explore the content of responses; thus, whether actions have been taken to address such concerns is unknown.

Our study reveals critical problems with the PFD system. Despite the legal requirement to respond to PFDs, there is no process for enforcing and auditing responses to ensure that necessary actions are being taken to prevent similar deaths. Others have suggested the need for sanctions to improve response rates and enforce Regulation 29 of The Coroners (Investigations) Regulations 2013 (9,37). But whether such sanctions would lead to increased actions for reducing preventable deaths has not been tested. In addition, the repeated concerns we identified have national importance for patient safety, which would benefit from creating an environment that encourages lessons to be shared and learnt. Building such an environment could be established using a quarterly summary of coroners’ reports, targeting key stakeholders, including policymakers, clinicians, and the public (39). Sharing concerns broadly may help disseminate lessons and allow locally relevant PFDs to have national benefits, facilitating changes by local organisations in response to other tragedies, rather than just reactions to their own. This may also require alterations to the content and recipients of PFDs to facilitate the learning of lessons elsewhere, such that they are also designed to provide general information to third parties.

Our research, alongside other studies (10,12,24,34,38–43), highlights that PFDs contain a wealth of important information that has merit for wide dissemination. However, crucial information in PFDs is often missing, incorrect, or inconsistent. Reporting of information could be improved using technologies that replace the current narrative system with an electronic form, providing specific fields to populate. This would reduce the time spent manually extracting data for research and improve the ability of the Preventable Deaths Tracker to analyse thousands of PFDs to generate summaries for dissemination.

Future research should explore the 1245 responses received from medicines-related PFDs to identify previously taken actions that could be shared across healthcare settings. Examining such responses would also identify gaps that have not yet been addressed. With this knowledge, future research could assess the impact and value of PFDs in changing clinical practice, which remains debated (10,12,24,34,38,39).

## Conclusions

One in five PFDs involved medicines and highlighted many areas of concern for patient care. Patients may benefit from changes to clinical practice surrounding medicines such as opioids and antidepressants, especially when related to problems with monitoring, record keeping, and communication. Patients and clinicians may also benefit from improvements to the PFD system, better reporting and resolution of concerning factors by organisations having the potential to reduce medicines-related deaths across England and Wales.

## Data Availability

All study materials, data, and statistical code is openly available via online repositories. The study protocol was preregistered on the Open Science Framework (OSF; https://doi.org/10.17605/OSF.IO/TX3CS); the code to generate the database and the Preventable Deaths Tracker is openly available via GitHub (https://github.com/georgiarichards/georgiarichards.github.io); individual Prevention of Future Deaths reports are available on the Courts and Tribunals Judiciary website (https://www.judiciary.uk/prevention-of-future-death-reports/); all other study materials are openly available via the OSF project page (https://doi.org/10.17605/OSF.IO/WQ7G5).

https://doi.org/10.17605/OSF.IO/TX3CS

## Data Availability

https://doi.org/10.17605/OSF.IO/TX3CS

## Authors and contributors

HSF updated the study protocol, carried out screening of PFDs, extracted remaining data, conducted all analyses, produced all figures and tables, and wrote the first draft of the manuscript. GCR conceptualised, designed, and initiated the study; provided supervisory support, and edited the first draft of the manuscript. REF and ARC contributed to study conceptualisation. CH and JKA provided supervisory support and oversight. All study authors read, contributed to, and approved the final manuscript. HSF is the guarantor. The corresponding author attests that all listed authors meet authorship criteria and that no others meeting the criteria have been omitted.

