## Supplement for "Preventable deaths involving medicines in England and Wales, 2013-22: a systematic case series of coroners’ reports"

**Supplement 1: Definition of a medicine**

Invoking previous definitions (1,2) we defined a medicine as a product ‘that contains a compound with proven biological effects… taken by or administered to a person or animal for one or more of the following reasons: (i) as a placebo; (ii) to prevent a disease; (iii) to make a diagnosis; (iv) to modify a physiological, biochemical, or anatomical function or abnormality; (vi) to replace a missing factor; (vii) to ameliorate a symptom; (viii) to treat a disease; and/or (ix) to induce anaesthesia’. Under (iv) we included drugs of abuse.

**Table S1:** Data fields extracted from Prevention of Future Deaths reports (PFDs) through the web scrape (left) or manually (right). The data were then presented in a tabular database for analysis.

| **Extracted Automatically** | **Extracted Manually** |
| --- | --- |
| Case reference number/url | Date of death(s) |
| Date of report | Inquest duration |
| Name of the deceased | Age of the deceased at death |
|  | Sex of the deceased |
|  | Relevant medical/social history of the deceased |
| Death category | Death setting(s) |
|  | Medical cause(s) of death |
|  | Coroner’s conclusion(s) |
|  | Class of medicine(s) involved in the death(s) |
|  | Details of the coroner’s concern(s) |
|  | Classification(s) of the coroner’s concern(s) |
|  | Action(s) proposed by the coroner |
|  | Classification(s) of action(s) proposed by the coroner |
| Those to whom the report was sent | Numbers and types of individuals or organizations to whom reports were sent |
|  | Date on which the response was due |
|  | Whether individuals or organizations addressed responded (yes/no) |
|  | Date of the response |
|  | Number of days underdue/overdue |

**Table S2**: Reporting rates of medicine-related Prevention of Future Deaths reports (PFDs) in England and Wales between July 2013 and 23 February 2022, normalized for year by calculating the mean number of reports published per day in the screened period

| **Year** | **Total PFD rate (reports/day)** | **Number of medicine-related PFDs** | **Medicine-related PFD rate (%)** |
| --- | --- | --- | --- |
| 2013 | 1.11 | 173 | 15.0 |
| 2014 | 1.52 | 555 | 15.1 |
| 2015 | 1.31 | 477 | 16.1 |
| 2016 | 1.28 | 470 | 14.0 |
| 2017 | 1.19 | 435 | 17.0 |
| 2018 | 1.14 | 415 | 20.7 |
| 2019 | 1.42 | 517 | 21.1 |
| 2020 | 0.86 | 313 | 26.2 |
| 2021 | 1.18 | 430 | 20.5 |
| 2022 | 1.06 | 52 | 23.1 |

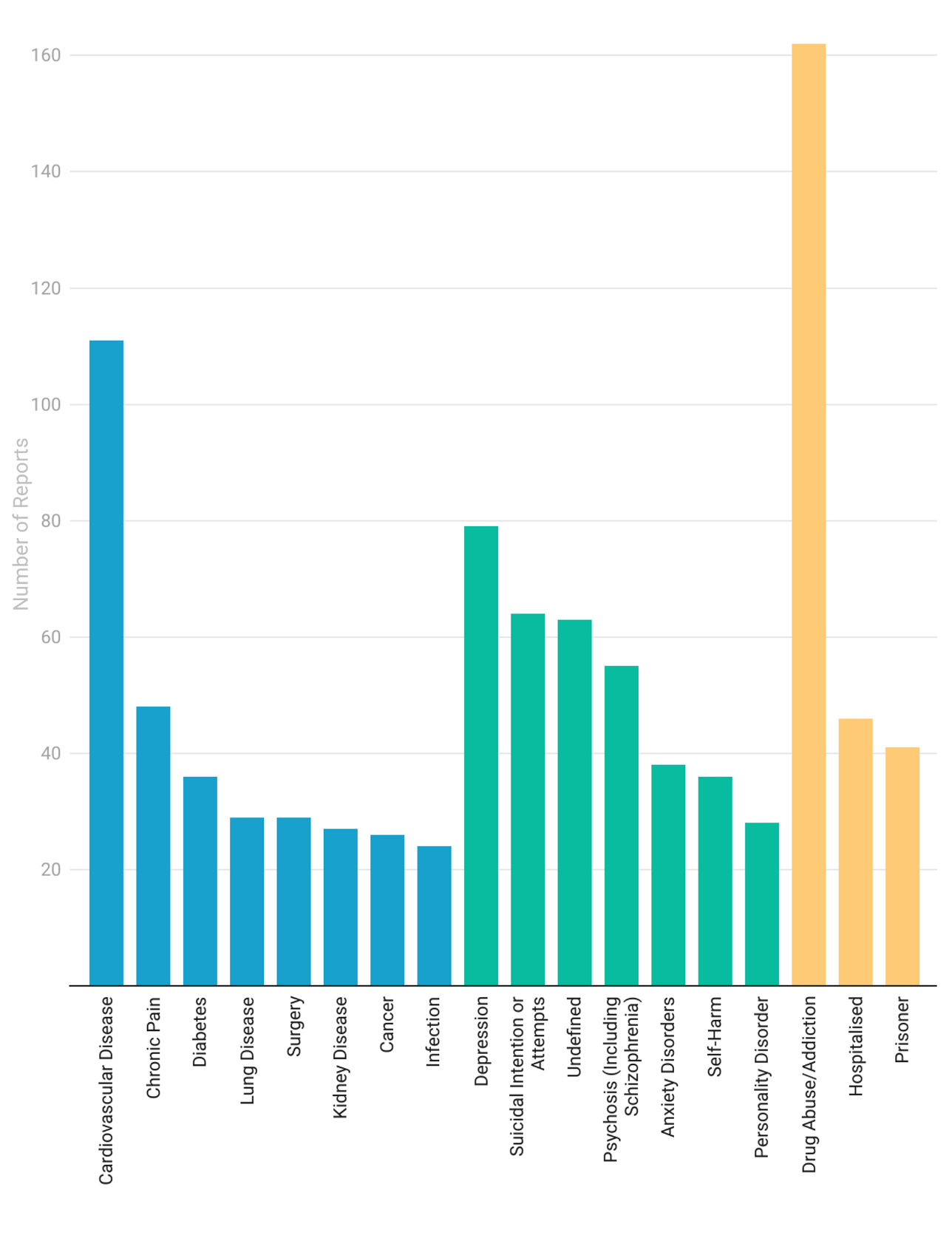

**Figure S1:** Frequency of types of case-relevant medical problems (blue), mental health problems (green) and social problems (yellow) of the deceased as reported by coroners in 704 Prevention of Future Death Reports (PFDs) involving medicines in England and Wales between July 2013 and 23 February 2022. Only common occurrences reported in >20 cases are presented in the figure; a complete list of extracted problems is provided in Table S3(A-C) in Supplement.

**Table S3A:** Reported past medical problems of the deceased mentioned by coroners in Prevention of Future Death Reports (PFDs) involving medicines in England and Wales between July 2013 and 23 February 2022

| **Condition** | **Number of PFD reports** |
| --- | --- |
| Cardiovascular Disease | 111 |
| Chronic Pain | 48 |
| Diabetes | 36 |
| Lung Disease | 29 |
| Surgery | 29 |
| Kidney Disease | 27 |
| Cancer | 26 |
| Infection | 24 |
| Epilepsy | 17 |
| Liver Disease | 17 |
| Movement Disorder | 14 |
| Undefined | 14 |
| Allergy | 13 |
| Asthma | 13 |
| Trauma | 13 |
| Stroke | 11 |
| Hormonal Imbalance | 10 |
| Obesity | 10 |
| Fall | 8 |
| Immune Dysfunction | 8 |
| Visual Issues | 7 |
| Frailty | 6 |
| Gastrointestinal Disease | 6 |
| Metabolic Disorder | 5 |
| Nutrient Deficit | 5 |
| Premature Birth | 4 |
| Arthritis | 3 |
| Down’s Syndrome | 3 |
| Skin Condition | 3 |
| Pregnancy | 2 |
| Total | 522 |

**Table S3B:** Reported past mental health problems of the deceased mentioned by coroners in Prevention of Future Death Reports (PFDs) involving medicines in England and Wales between July 2013 and 23 February 2022.

| **Condition** | **Number of PFD reports** |
| --- | --- |
| Depression | 79 |
| Suicidal Intention or Attempts | 64 |
| Undefined | 63 |
| Psychosis (Including Schizophrenia) | 55 |
| Anxiety Disorders | 38 |
| Self-Harm | 36 |
| Personality Disorder | 28 |
| Dementia | 15 |
| Bipolar Affective Disorder | 14 |
| ADHD | 10 |
| ASD | 10 |
| PTSD | 9 |
| Behavioural Disorder | 4 |
| Parkinson’s Disease | 4 |
| Eating Disorder | 3 |
| Mood Disorders | 3 |
| OCD | 3 |
| Emotional Disorder | 2 |
| Phobia | 2 |
| Total | 442 |

*ADHD*: Attention Deficit Hyperactive Disorder; *ASD*: Autistic Spectrum Disorder; *PTSD*: Post-Traumatic Stress Disorder; *OCD*: Obsessive-Compulsive Disorder.

**Table S3C:** Reported past social problems of the deceased mentioned by coroners in Prevention of Future Death Reports (PFDs) involving medicines in England and Wales between July 2013 and 23 February 2022.

| **Category** | **Number of PFD Reports** |
| --- | --- |
| Drug Abuse/Addiction | 162 |
| Hospitalized (Voluntarily or on Mental Health Grounds) | 46 |
| Prisoner | 41 |
| Past Overdose | 19 |
| Poor Engagement with Healthcare/Community Services | 12 |
| Relationship Problems | 12 |
| Care Home | 11 |
| Homelessness | 11 |
| Student | 8 |
| Death in Family | 7 |
| Having Financial Problems | 7 |
| Abuse Victim | 5 |
| Assault Victim | 5 |
| Isolated Individual | 5 |
| Social Problems | 5 |
| Drug Trial Participant | 4 |
| Good Engagement with Healthcare/Community Services | 4 |
| Previous Child Death | 4 |
| Event Attendee | 3 |
| DNAR Order | 2 |
| Fall | 2 |
| Insufficient Care | 2 |
| Professional Issues | 2 |
| Asylum Seeker | 1 |
| Foster Child | 1 |
| Missing Person | 1 |
| Pilot | 1 |
| Total | 383 |

*DNAR*: Do Not Attempt Resuscitation.

**Table S4**: Distribution of Prevention of Future Deaths reports (PFDs) in England and Wales between July 2013 and 23 February 2022 by coroner’s area. Total number of medicine-related PFDs was used to generate a decile ranking of areas by frequency, and then used to calculate the rate of medicine-related PFDs from the total produced from each area for an additional ranking by decile.

| **Coroners’ Area** | **Total PFDs** | **Medicine-Related PFDs** | **Decile Rank (Number of Medicine-Related Reports)** | **Rate of Medicine-Related PFDs** | **Decile Rank (Rate)** |
| --- | --- | --- | --- | --- | --- |
| Avon | 71 | 12 | 8 | 0.169 | 6 |
| Bedfordshire & Luton | 58 | 12 | 8 | 0.207 | 7 |
| Berkshire | 33 | 5 | 5 | 0.152 | 5 |
| Birmingham & Solihull | 114 | 29 | 10 | 0.254 | 9 |
| Blackpool & Fylde | 35 | 9 | 7 | 0.257 | 9 |
| Brighton & Hove | 61 | 8 | 6 | 0.131 | 4 |
| Buckinghamshire | 17 | 3 | 3 | 0.176 | 6 |
| Cambridgeshire & Peterborough | 44 | 8 | 6 | 0.182 | 6 |
| Carmarthenshire & Pembrokeshire | 4 | 0 | 1 | 0.000 | 1 |
| Ceredigon | 1 | 0 | 1 | 0.000 | 1 |
| Cheshire | 22 | 2 | 1 | 0.091 | 2 |
| Cornwall & The Isles of Scilly | 71 | 14 | 9 | 0.197 | 7 |
| County Durham & Darlington | 63 | 11 | 7 | 0.175 | 6 |
| Coventry | 20 | 6 | 6 | 0.300 | 10 |
| Cumbria | 51 | 6 | 6 | 0.118 | 3 |
| Derby & Derbyshire | 29 | 4 | 4 | 0.138 | 4 |
| Dorset | 38 | 11 | 7 | 0.289 | 10 |
| East Riding of Yorkshire & Kingston-Upon-Hull | 10 | 2 | 3 | 0.200 | 7 |
| East Sussex | 11 | 3 | 3 | 0.273 | 10 |
| Essex | 52 | 13 | 8 | 0.250 | 9 |
| Exeter & Greater Devon | 43 | 15 | 9 | 0.349 | 10 |
| Gateshead & South Tyneside | 7 | 1 | 2 | 0.143 | 4 |
| Gloucestershire | 32 | 5 | 5 | 0.156 | 5 |
| Greater Manchester North | 95 | 29 | 10 | 0.305 | 10 |
| Greater Manchester South | 316 | 61 | 10 | 0.193 | 7 |
| Greater Manchester West | 103 | 23 | 10 | 0.223 | 8 |
| Gwent | 27 | 3 | 3 | 0.111 | 3 |
| Hampshire, Portsmouth & Southampton | 64 | 14 | 9 | 0.219 | 8 |
| Herefordshire | 3 | 1 | 2 | 0.333 | 10 |
| Hertfordshire | 28 | 4 | 4 | 0.143 | 4 |
| Isle of Wight | 19 | 5 | 5 | 0.263 | 9 |
| Kent Central & South East | 23 | 3 | 3 | 0.130 | 4 |
| Lancashire with Blackburn & Darwen | 44 | 5 | 5 | 0.114 | 3 |
| Leicester & South Leicestershire | 60 | 12 | 8 | 0.200 | 7 |
| Lincolnshire | 48 | 8 | 6 | 0.167 | 5 |
| Liverpool & Wirral | 44 | 3 | 3 | 0.068 | 2 |
| London City | 11 | 3 | 3 | 0.273 | 9 |
| London East | 91 | 19 | 10 | 0.209 | 7 |
| London Inner North | 193 | 25 | 10 | 0.130 | 4 |
| London Inner South | 110 | 20 | 10 | 0.182 | 6 |
| London Inner West | 54 | 12 | 8 | 0.222 | 8 |
| London North | 52 | 9 | 7 | 0.173 | 6 |
| London South | 34 | 5 | 5 | 0.147 | 5 |
| London West | 38 | 9 | 7 | 0.237 | 9 |
| Manchester City | 53 | 15 | 9 | 0.283 | 10 |
| Mid Kent & Medway | 47 | 5 | 5 | 0.106 | 3 |
| Milton Keynes | 53 | 5 | 5 | 0.094 | 3 |
| Newcastle Upon Tyne | 20 | 2 | 3 | 0.100 | 3 |
| Norfolk | 80 | 9 | 7 | 0.113 | 3 |
| North East Kent | 11 | 2 | 3 | 0.182 | 6 |
| North Lincolnshire & Grimsby | 4 | 0 | 1 | 0.000 | 1 |
| North Northumberland | 11 | 0 | 1 | 0.000 | 1 |
| North Tyneside | 0 | 0 | 1 | 0.000 | 1 |
| North Wales (East & Central) | 62 | 8 | 6 | 0.129 | 4 |
| North West Kent | 7 | 0 | 1 | 0.000 | 1 |
| North West Wales | 11 | 0 | 1 | 0.000 | 1 |
| North Yorkshire (Eastern) | 1 | 0 | 1 | 0.000 | 1 |
| North Yorkshire (Western) | 9 | 0 | 1 | 0.000 | 1 |
| Northamptonshire | 23 | 4 | 4 | 0.174 | 6 |
| Nottinghamshire & Nottingham | 79 | 17 | 9 | 0.215 | 8 |
| Oxfordshire | 25 | 3 | 3 | 0.120 | 4 |
| Plymouth, Torbay and South Devon | 45 | 7 | 6 | 0.156 | 5 |
| Rutland & North Leicestershire | 7 | 0 | 1 | 0.000 | 1 |
| Sefton, St. Helens and Knowsley | 1 | 0 | 1 | 0.000 | 1 |
| Shropshire, Telford & Wrekin | 22 | 3 | 3 | 0.136 | 4 |
| Somerset | 13 | 1 | 2 | 0.077 | 2 |
| South Northumberland | 1 | 0 | 1 | 0.000 | 1 |
| South Wales Central | 101 | 22 | 10 | 0.218 | 8 |
| South Yorkshire (East) | 44 | 6 | 6 | 0.136 | 4 |
| South Yorkshire (West) | 54 | 4 | 4 | 0.074 | 2 |
| Staffordshire South | 60 | 13 | 8 | 0.217 | 8 |
| Stoke-on-Trent & North Staffordshire | 40 | 10 | 7 | 0.250 | 9 |
| Suffolk | 37 | 9 | 7 | 0.243 | 9 |
| Sunderland | 35 | 0 | 1 | 0.000 | 1 |
| Surrey | 105 | 24 | 10 | 0.229 | 8 |
| Swansea, Neath & Port Talbot | 17 | 5 | 5 | 0.294 | 10 |
| Teesside & Hartlepool | 13 | 2 | 3 | 0.154 | 5 |
| The Black Country Jurisdiction | 74 | 16 | 9 | 0.216 | 8 |
| Warwickshire | 28 | 3 | 3 | 0.107 | 3 |
| West Sussex | 67 | 13 | 8 | 0.194 | 7 |
| West Yorkshire (Eastern) | 84 | 13 | 8 | 0.155 | 5 |
| West Yorkshire (Western) | 64 | 12 | 8 | 0.188 | 7 |
| Wiltshire & Swindon | 45 | 11 | 7 | 0.244 | 9 |
| Worcestershire | 38 | 7 | 6 | 0.184 | 7 |
| York | 3 | 1 | 2 | 0.333 | 10 |

**Table S5:** Number and rate of medicines reported in Prevention of Future Deaths reports (PFDs) in England and Wales between July 2013 and 23 February 2022

| **Number of medicines** | **Number of PFDs (%)** |
| --- | --- |
| 1 | 454 (64.5) |
| 2 | 145 (20.6) |
| 3 | 57 (8.1) |
| 4 | 23 (3.3) |
| 5 | 10 (1.4) |
| 6 | 6 (0.9) |
| 7 | 1 (0.1) |
| 8 | 4 (0.6) |
| 9 | 1 (0.1) |
| 10 | 1 (0.1) |
| 11 | 0 |
| 12 | 2 (0.3) |

**Table S6**: All drug classes reported in included Prevention of Future Deaths reports (PFDs) in England and Wales between July 2013 and 23 February 2022, reported by frequency

| **Drug Class** | **Cases** |
| --- | --- |
| Opioid Analgesics | 257 |
| Antidepressant Drugs | 114 |
| Hypnotics | 108 |
| Anticoagulants | 85 |
| Antipsychotic Drugs | 54 |
| Antibiotics | 47 |
| Cocaine | 47 |
| Antiepileptic Drugs | 43 |
| Cannabinoids | 40 |
| Non-Opioid Analgesics | 28 |
| Hallucinogenic Substances | 25 |
| Stimulants | 24 |
| Non-Steroidal Anti-Inflammatory Drugs | 19 |
| Volatile Substances and Gases, Not Otherwise Covered | 17 |
| Insulin | 14 |
| Beta-Blockers | 13 |
| Steroids | 12 |
| Antihistamines | 11 |
| Drugs for Malignant Disease and Immunosuppression | 10 |
| Supplements | 9 |
| Emollients | 8 |
| General Anaesthetics | 8 |
| Proton Pump Inhibitors | 8 |
| Sodium Nitrates | 8 |
| Drugs Affecting the Renin-Angiotensin System | 6 |
| Ketamine | 6 |
| Lithium Salts | 6 |
| Poisoning Antidotes | 6 |
| Sympathomimetics | 6 |
| Antiarrhythmic Drugs | 5 |
| Antimuscarinic Drugs | 5 |
| Pure Immunosuppressants | 5 |
| Uncoupling agents | 4 |
| Antihypertensive Drugs | 3 |
| Fibrinolytics | 3 |
| Local Anaesthetics | 3 |
| X-Ray Contrast Media | 3 |
| Antidiabetic Drugs | 2 |
| Coagulation Proteins | 2 |
| Diuretics | 2 |
| Monoclonal Antibodies | 2 |
| Neuromuscular Blocking Drugs | 2 |
| Prostaglandin Analogues | 2 |
| Statins | 2 |
| Xanthine Oxidase inhibitors | 2 |
| Antipropulsives | 1 |
| Antiseptic | 1 |
| Carbonic Anhydrase inhibitors | 1 |
| Dopamine Antagonists | 1 |
| Laxatives | 1 |
| Organophosphates | 1 |
| Vaccines | 1 |
| Vasopressin and Analogues | 1 |
| Undefined | 78 |

Supplement 2: Illustrative Examples of Cases and Concerns

a) ***Regulation: Inadequate drug regulations enabled access to 2,4-dinitrophenol tablets over the internet (2020-0144)***(3)

The deceased was admitted to hospital and confirmed taking an overdose of 2,4-DNP tablets. Despite appropriate treatment, he died later the same day. The coroner believed that he had used the internet to acquire DNP for weight loss, attempting to improve his body image. The coroner was concerned that, although DNP was banned in 1952(4), it was still accessible over the ‘dark web’.

b) ***Patient Safety: Medicine administration errors led to excessive bleeding, which was inadequately managed (2018-0146)***(5)

The deceased was found bleeding through an arteriovenous graft, having been given warfarin by mistake over the previous few days. Although he lived in a care home, staff made no attempt to halt the bleed until paramedics arrived. The coroner was concerned that unprescribed warfarin had been repeatedly administered to the deceased, facilitating the bleed. Furthermore, when the bleed occurred, care home staff lacked patient information and the ability to deal with the emergency. This indicated inadequate training and poor access to medical records, which might have helped the ambulance crew prevent the death.

c) ***Resources: Lack of protocol regarding the removal of medicines from a deceased individual contributed to an impulsive suicide (2020-0077)***(6)

The deceased moved to a new address and accessed prescription oral morphine left here after the death of the previous resident. In combination with midazolam, paracetamol, and alcohol, this precipitated his impulsive suicide. The coroner was concerned by the lack of arrangements or guidance regarding the removal of remaining prescriptions after a patient’s death.

d) ***Failure to Follow Necessary Tasks/Protocols/Jobs: Administration of thrombolytic agents in contrast to NICE guidelines led to death (2015-0054)***(7)

The deceased had previously suffered from a stroke and was being managed in hospital, but no venous thromboembolism (VTE) assessment was carried out during this period. Consequently, NICE guidelines to use mechanical anti-thrombotic devices instead of pharmacotherapy was ignored, with the subsequent administration of thromboembolytics in response to a pulmonary embolus (PE), leading to catastrophic bleeding and death.

The coroner was concerned that the decision not to provide a VTE assessment was unjustified, and that staff failed to follow NICE guidance during the treatment of the PE.

e) ***Communication: Poor communication between organizations left a vulnerable individual unsupported, precipitating an intentional venlafaxine overdose (2018-0283)***(8)

After a previous suicide attempt and hospitalization, the deceased was referred to a crisis team by hospital staff, who then referred him again to mental health service counselling. Responsibility was placed on the deceased to organize this transfer, and the crisis team did not communicate with mental health services or follow up he patient. This meant that the deceased lacked support, facilitating his suicide later that month.

The coroner was concerned that excessive responsibility had been placed on a vulnerable patient to organize their own continued care, and that there was no system of communication between the hospital staff and mental health care providers, which could have prompted follow-up or more active transfer.

f) ***Education & Training: Inadequate training prevented the police from properly responding to a case of excited delirium brought about by use of cocaine (2013-0352)***(9)

The deceased had returned from a trip, during which he had consumed cocaine, alcohol, and illicit Valium, leading to erratic and alarming behaviour, which prompted the public to contact emergency services. The police apprehended the deceased and restrained him, leading to further agitation. The deceased suffered a cardiac arrest while in an ambulance and could not be resuscitated.

Although criticising neither the police officers nor paramedics, the coroner was concerned about the lack of training provided to emergency service staff regarding the state of excited delirium. The coroner acknowledged that such training had since been implemented by the local police, but wanted to determine whether the change has been disseminated nationally.

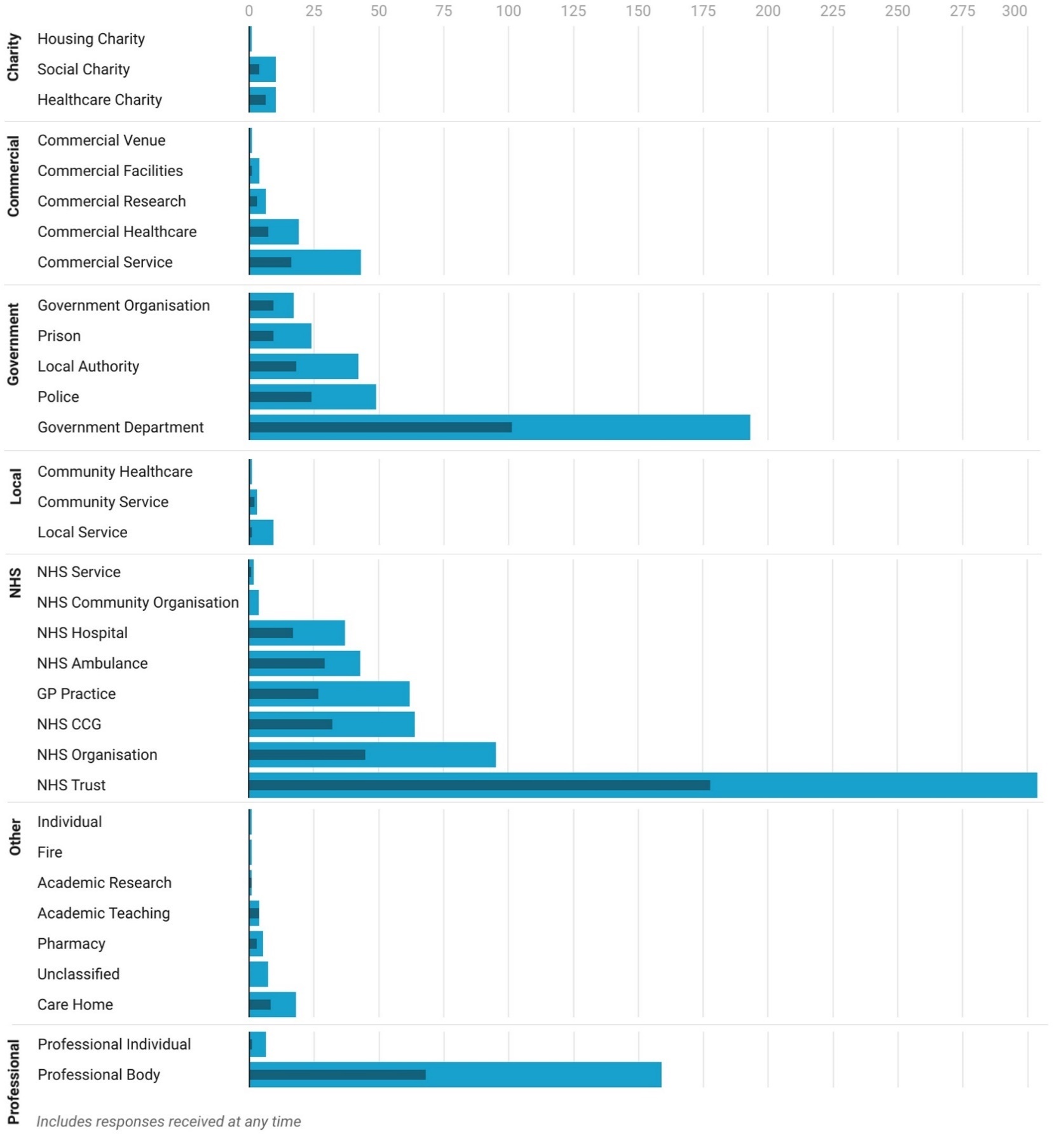
**Figure S2**: Total number of medicine-related Prevention of Future Death reports (PFDs) in England and Wales published between July 2013 and 23 February 2022 sent to different categories of organizations/individuals (light blue), and responses received (dark blue). *CCG = Clinical Commissioning Group; GP = General Practice; Undefined = addressee not named by coroner.*

**Table S7**: Individuals/Organizations to which the 704 included medicines-related Prevention of Future Death reports (PFDs) published between July 2013 and 23 February 2022 were sent, and the frequency of their different responses rates

| **Organization/Individual** | **Type of Organization** | **Early (>7)** | **On Time**  **(-7>t>7)** | **Late**  **(<-7)** | **Overdue** | **Total** |
| --- | --- | --- | --- | --- | --- | --- |
| ------------------------------------------------------- | Academic Research | 0 | 1 | 0 | 0 | 1 |
| Drug Misuse and Novel Psychoactive Substance Unit, University of Hertfordshire | Academic Research |  | 1 |  |  | 1 |
| ------------------------------------------------------- | Academic Teaching | 0 | 3 | 1 | 0 | 4 |
| St John's College, University of Oxford | Academic Teaching |  |  | 1 |  | 1 |
| University of Exeter | Academic Teaching |  | 1 |  |  | 1 |
| University of South Wales | Academic Teaching |  | 1 |  |  | 1 |
| Warwick Medical School | Academic Teaching |  | 1 |  |  | 1 |
| ------------------------------------------------------- | Care Home | 2 | 4 | 2 | 10 | 18 |
| Adbolton Hall Nursing Home | Care Home |  |  | 1 |  | 1 |
| Barchester Homes | Care Home |  | 1 |  |  | 1 |
| BUPA Care Homes | Care Home |  |  |  | 1 | 1 |
| Care UK | Care Home | 2 |  |  | 3 | 5 |
| Castlehill Specialist Care Centre | Care Home |  |  | 1 |  | 1 |
| Dovecote Lodge | Care Home |  |  |  | 1 | 1 |
| Grassy Meadow Care Centre | Care Home |  | 1 |  |  | 1 |
| HC-One | Care Home |  | 1 |  |  | 1 |
| Jeesal Residential Care Services | Care Home |  | 1 |  |  | 1 |
| Lapal House & Lodge Care Home | Care Home |  |  |  | 1 | 1 |
| Oak Lodge Care Home | Care Home |  |  |  | 1 | 1 |
| Passmonds Care Home | Care Home |  |  |  | 1 | 1 |
| Pindy Enterprises Limited | Care Home |  |  |  | 1 | 1 |
| Riverview | Care Home |  |  |  | 1 | 1 |
| ------------------------------------------------------- | Commercial Facilities | 0 | 1 | 0 | 3 | 4 |
| Bannatyne Group | Commercial Facilities |  | 1 |  |  | 1 |
| Jurys Inn Birmingham | Commercial Facilities |  |  |  | 1 | 1 |
| Trent and Dove Social Housing | Commercial Facilities |  |  |  | 1 | 1 |
| Worthing Homes | Commercial Facilities |  |  |  | 1 | 1 |
| ------------------------------------------------------- | Commercial Healthcare | 1 | 4 | 2 | 12 | 19 |
| 1st for Health International Ltd | Commercial Healthcare |  |  |  | 1 | 1 |
| Advanced Medical Priority Dispatch | Commercial Healthcare |  |  |  | 1 | 1 |
| Alliance Pharmaceutical | Commercial Healthcare |  |  | 1 |  | 1 |
| Appello | Commercial Healthcare |  |  |  | 1 | 1 |
| Central Medical Services | Commercial Healthcare |  |  |  | 1 | 1 |
| Comfort Call | Commercial Healthcare |  | 1 |  |  | 1 |
| CRG Lead Commissioner | Commercial Healthcare |  |  |  | 1 | 1 |
| First Class Care | Commercial Healthcare |  |  |  | 1 | 1 |
| Livewell South West | Commercial Healthcare |  |  |  | 2 | 2 |
| Manchester Medical Services | Commercial Healthcare |  |  |  | 1 | 1 |
| Masta Limited | Commercial Healthcare |  | 1 |  |  | 1 |
| Nestor Primecare | Commercial Healthcare |  |  |  | 1 | 1 |
| Next Stage | Commercial Healthcare | 1 |  |  |  | 1 |
| PJ Care | Commercial Healthcare |  |  |  | 1 | 1 |
| Priory Group | Commercial Healthcare |  | 1 | 1 |  | 2 |
| Reckitt Benckisher Healthcare (UK) Ltd | Commercial Healthcare |  | 1 |  |  | 1 |
| Tunstall Response | Commercial Healthcare |  |  |  | 1 | 1 |
| ------------------------------------------------------- | Commercial Research | 0 | 2 | 1 | 3 | 6 |
| Amgen Limited | Commercial Research |  |  |  | 1 | 1 |
| Bayer Plc | Commercial Research |  | 1 |  |  | 1 |
| Boehringer Ingelheim Ltd | Commercial Research |  | 1 |  |  | 1 |
| Diprobase Bayer Public Limited | Commercial Research |  |  |  | 1 | 1 |
| Takeda Pharmaceutical Company | Commercial Research |  |  | 1 |  | 1 |
| UCB Pharma | Commercial Research |  |  |  | 1 | 1 |
| ------------------------------------------------------- | Commercial Service | 3 | 7 | 6 | 27 | 43 |
| 700 Club | Commercial Service |  |  | 1 |  | 1 |
| Cambian Group | Commercial Service |  |  | 1 |  | 1 |
| CAMIS | Commercial Service |  |  |  | 1 | 1 |
| Capita Business Services Ltd | Commercial Service |  | 1 |  |  | 1 |
| Carlsberg Supply Co Ltd | Commercial Service |  | 1 |  |  | 1 |
| Ebay UK Ltd | Commercial Service |  | 1 |  | 1 | 2 |
| EMIS | Commercial Service |  |  |  | 1 | 1 |
| Farleys Solicitors | Commercial Service |  |  |  | 1 | 1 |
| Festival Republic Ltd and Live Nation Entertainment PLC | Commercial Service |  |  | 1 |  | 1 |
| First Response Group | Commercial Service |  |  |  | 1 | 1 |
| G4S | Commercial Service |  | 1 | 1 | 3 | 5 |
| Guest Medical | Commercial Service |  |  |  | 1 | 1 |
| H R Healthcare Limited | Commercial Service |  |  |  | 1 | 1 |
| Haulage Contractors Limited | Commercial Service |  |  |  | 1 | 1 |
| Hoults Limited | Commercial Service |  |  |  | 1 | 1 |
| Informa Healthcare | Commercial Service |  |  |  | 1 | 1 |
| Jigsaw Homes Group | Commercial Service |  |  |  | 1 | 1 |
| Lifeskills Medical UK | Commercial Service |  |  |  | 1 | 1 |
| Medtronic | Commercial Service |  |  |  | 1 | 1 |
| Metalchem Ltd | Commercial Service | 1 |  |  |  | 1 |
| Mitie Group | Commercial Service |  |  |  | 3 | 3 |
| Oasis Recovery Communities | Commercial Service |  |  | 1 |  | 1 |
| Proprietary Association | Commercial Service |  | 1 |  |  | 1 |
| Radcliffe Investment Properties | Commercial Service | 1 |  |  |  | 1 |
| Roche Diagnostics Limited | Commercial Service |  | 1 |  |  | 1 |
| Royal Mail | Commercial Service | 1 |  |  |  | 1 |
| Serco | Commercial Service |  |  |  | 1 | 1 |
| Shindig Events Limited | Commercial Service |  |  |  | 1 | 1 |
| Sodexo Justice Services | Commercial Service |  |  |  | 1 | 1 |
| StockXS Ltd | Commercial Service |  |  |  | 1 | 1 |
| SystemOne TPP Ltd | Commercial Service |  |  |  | 1 | 1 |
| Teva UK | Commercial Service |  |  | 1 |  | 1 |
| Treatment Direct Ltd | Commercial Service |  |  |  | 1 | 1 |
| TUI UK & Ireland | Commercial Service |  |  |  | 1 | 1 |
| Wellsky | Commercial Service |  |  |  | 1 | 1 |
| British Airways | Commercial Services |  | 1 |  |  | 1 |
| ------------------------------------------------------- | Commercial Venue | 0 | 0 | 0 | 1 | 1 |
| Egg London Nightclub | Commercial Venue |  |  |  | 1 | 1 |
| ------------------------------------------------------- | Community Healthcare | 0 | 0 | 0 | 1 | 1 |
| Lincolnshire Community Health Services | Community Healthcare |  |  |  | 1 | 1 |
| ------------------------------------------------------- | Community Service | 1 | 1 | 0 | 1 | 3 |
| Adult Safeguarding Civic Service | Community Service |  | 1 |  |  | 1 |
| Adult Social Services | Community Service | 1 |  |  |  | 1 |
| Wandsworth Consortium Drug and Alcohol Services | Community Service |  |  |  | 1 | 1 |
| ------------------------------------------------------- | Fire | 0 | 0 | 0 | 1 | 1 |
| Staffordshire Fire & Rescue Service | Fire |  |  |  | 1 | 1 |
| ------------------------------------------------------- | Government Department | 15 | 34 | 52 | 92 | 193 |
| Department for Business, Innovation & Skills (Consumer Safety) | Government Department | 1 |  |  | 1 | 2 |
| Department for Digital, Culture, Media & Sport | Government Department |  |  |  | 2 | 2 |
| Department for Transport | Government Department |  |  | 1 | 1 | 2 |
| Department for Work and Pensions | Government Department |  | 2 | 1 |  | 3 |
| Department of Health & Social Care | Government Department | 5 | 10 | 27 | 25 | 67 |
| DVLA | Government Department |  |  |  | 2 | 2 |
| FCO | Government Department | 1 |  |  |  | 1 |
| Food Standards Agency | Government Department |  | 1 |  |  | 1 |
| Health & Safety Executive | Government Department | 1 |  |  |  | 1 |
| Health Education England | Government Department |  |  |  | 1 | 1 |
| HM Prison and Probation Service | Government Department |  | 5 | 4 | 6 | 15 |
| Home Office | Government Department | 1 | 4 | 4 | 12 | 21 |
| Minister for Education | Government Department |  |  |  | 1 | 1 |
| Minister for Local Government | Government Department |  |  |  | 1 | 1 |
| Minister for Policing, Fire and Criminal Justice | Government Department |  | 2 |  | 3 | 5 |
| Minister of Health | Government Department |  | 1 | 2 |  | 3 |
| Minister of State for Care and Support | Government Department |  |  | 1 |  | 1 |
| Minister of State for Crime Prevention | Government Department |  |  |  | 3 | 3 |
| Minister of State for Mental Health, Suicide Prevention and Patient Safety | Government Department |  | 1 | 1 | 3 | 5 |
| Minister of State for Security | Government Department |  |  |  | 1 | 1 |
| Ministry of Defence, Welfare and Veterans | Government Department |  | 1 |  |  | 1 |
| MOJ | Government Department | 1 |  |  | 10 | 11 |
| National Crime Agency | Government Department |  |  |  | 1 | 1 |
| NOMS | Government Department | 1 | 1 | 2 | 3 | 7 |
| Secretary of State for Education | Government Department |  |  |  | 1 | 1 |
| Secretary of State for Health | Government Department | 4 | 5 | 8 | 11 | 28 |
| Secretary of State for Justice | Government Department |  |  |  | 1 | 1 |
| Welsh Minister for Health | Government Department |  | 1 | 1 | 3 | 5 |
| ------------------------------------------------------- | Government Organization | 6 | 2 | 1 | 8 | 17 |
| Advisory Council on the Misuse of Drugs | Government Organization |  |  | 1 | 6 | 7 |
| Highways England | Government Organization |  | 1 |  |  | 1 |
| HSE | Government Organization | 1 |  |  |  | 1 |
| Network Rail | Government Organization | 1 |  |  |  | 1 |
| Police and Prisons Ombudsman | Government Organization |  |  |  | 1 | 1 |
| Public Health England | Government Organization | 4 | 1 |  | 1 | 6 |
| ------------------------------------------------------- | GP Practice | 12 | 10 | 5 | 35 | 62 |
| Alexander House Health Centre | GP Practice | 1 |  |  |  | 1 |
| Axminster Medical Practice | GP Practice | 1 |  |  |  | 1 |
| Bexley Medical Group | GP Practice | 1 |  |  |  | 1 |
| Black Country Family Practice | GP Practice |  |  |  | 1 | 1 |
| Bodmin Road Health Centre | GP Practice | 1 |  |  |  | 1 |
| Brace Street Health Centre | GP Practice |  | 1 |  |  | 1 |
| Broadgate General Practice | GP Practice |  |  |  | 1 | 1 |
| Browning Street Surgery | GP Practice |  | 1 |  |  | 1 |
| Castlefields Health Centre | GP Practice |  |  |  | 1 | 1 |
| Churchgate Surgery | GP Practice |  |  | 1 |  | 1 |
| Cornerstone Family Practice | GP Practice |  |  |  | 1 | 1 |
| Cricket Green Medical Practice | GP Practice |  |  | 1 |  | 1 |
| Darwin medical Practice | GP Practice | 1 |  |  |  | 1 |
| Delamere Medical Practice | GP Practice |  |  |  | 1 | 1 |
| Dower House Surgery | GP Practice |  |  |  | 1 | 1 |
| Eltham Park Surgery | GP Practice |  |  |  | 1 | 1 |
| Fern House Surgery | GP Practice |  |  |  | 1 | 1 |
| Fremington Medical Centre | GP Practice |  | 1 |  |  | 1 |
| Glenroyd Medical Practice | GP Practice |  |  |  | 1 | 1 |
| GP Practice Orchard Surgery | GP Practice |  |  |  | 2 | 2 |
| Grosvenor Medical Centre | GP Practice | 1 |  |  |  | 1 |
| HAZELMERE MEDICAL CENTRE | GP Practice |  | 1 |  |  | 1 |
| Heaton Moor Medical Centre | GP Practice |  |  |  | 3 | 3 |
| Heaton Norris Health Centre | GP Practice | 1 |  |  |  | 1 |
| Hopwood House Medical Practice | GP Practice | 1 |  |  |  | 1 |
| Ivy Grove Surgery | GP Practice |  |  |  | 1 | 1 |
| King Street Medical Practice | GP Practice |  |  |  | 1 | 1 |
| Knoll Surgery Partnership | GP Practice |  |  |  | 1 | 1 |
| Langley Health Centre | GP Practice |  |  |  | 1 | 1 |
| Lawn Medical Practice | GP Practice |  |  |  | 1 | 1 |
| Legal Counsel and Company Secretary The Practice | GP Practice |  |  |  | 1 | 1 |
| Lisson Grove Health Centre | GP Practice |  |  |  | 1 | 1 |
| Lodge Road Surgery | GP Practice |  |  |  | 1 | 1 |
| Long Furlong Medical Centre | GP Practice |  |  |  | 1 | 1 |
| Longshoot Health Centre | GP Practice |  |  |  | 1 | 1 |
| Manor Field Surgery | GP Practice |  | 1 |  |  | 1 |
| Medical Centre Stalybridge | GP Practice | 1 |  |  |  | 1 |
| Moat Surgery | GP Practice | 1 |  |  |  | 1 |
| New Court Surgery | GP Practice |  |  | 1 |  | 1 |
| Newbury Park Health Centre | GP Practice |  |  |  | 1 | 1 |
| Newgate Medical Group | GP Practice |  |  |  | 1 | 1 |
| North Laine Medical Centre | GP Practice |  |  |  | 1 | 1 |
| Northfield Medical Centre | GP Practice |  | 1 |  |  | 1 |
| Park View Group Practice | GP Practice |  |  |  | 1 | 1 |
| Practice Rose House | GP Practice |  |  |  | 1 | 1 |
| Princess Street Group Practice | GP Practice |  |  |  | 1 | 1 |
| Richmond Medical Centre | GP Practice |  |  |  | 1 | 1 |
| Rookery Medical Centre | GP Practice |  |  |  | 1 | 1 |
| Roundwell Medical Centre | GP Practice |  | 1 |  |  | 1 |
| Rush Green Medical Centre | GP Practice |  |  |  | 1 | 1 |
| Seaton and Colyton Medical Practice | GP Practice | 1 |  |  |  | 1 |
| St Chads Medical Practice | GP Practice |  | 1 |  |  | 1 |
| Tettenhall Medical Practice | GP Practice |  |  | 1 |  | 1 |
| Tredegar Practice | GP Practice |  |  |  | 1 | 1 |
| Uplands Medical Practice | GP Practice |  |  | 1 |  | 1 |
| Village Medical Centre | GP Practice |  |  |  | 1 | 1 |
| Wells Road Surgery | GP Practice | 1 |  |  |  | 1 |
| West Timperley Medical Centre | GP Practice |  | 1 |  |  | 1 |
| Woodley Centre Surgery | GP Practice |  | 1 |  |  | 1 |
| ------------------------------------------------------- | Healthcare Charity | 3 | 3 | 0 | 4 | 10 |
| Addiction Dependency Solutions | Healthcare Charity | 1 |  |  |  | 1 |
| CRI | Healthcare Charity |  |  |  | 1 | 1 |
| Free the Way | Healthcare Charity | 1 |  |  |  | 1 |
| The Forward Trust | Healthcare Charity |  |  |  | 1 | 1 |
| Humankind | Healthcare Charity |  | 1 |  |  | 1 |
| IC24 | Healthcare Charity |  | 1 |  |  | 1 |
| Locala | Healthcare Charity |  | 1 |  |  | 1 |
| MacMillan Cancer Care | Healthcare Charity |  |  |  | 1 | 1 |
| St John Ambulance | Healthcare Charity | 1 |  |  |  | 1 |
| WISH | Healthcare Charity |  |  |  | 1 | 1 |
| ------------------------------------------------------- | Housing Charity | 0 | 0 | 0 | 1 | 1 |
| Adullam Homes Housing Association | Housing Charity |  |  |  | 1 | 1 |
| ------------------------------------------------------- | Individual | 0 | 0 | 0 | 1 | 1 |
| Family | Individual |  |  |  | 1 | 1 |
| ------------------------------------------------------- | Local Authority | 2 | 13 | 3 | 24 | 42 |
| Banes Highways | Local Authority |  |  |  | 1 | 1 |
| Banes Park and Services | Local Authority |  |  |  | 1 | 1 |
| Birmingham City Council | Local Authority |  |  |  | 1 | 1 |
| Blaenau Gwent County Borough Council | Local Authority |  | 1 |  |  | 1 |
| Brighton & Hove City Council | Local Authority |  |  | 1 |  | 1 |
| Brighton and Hove Director of Housing | Local Authority |  |  |  | 1 | 1 |
| Brighton and Hove Health and Adult Social Care | Local Authority |  |  |  | 1 | 1 |
| Brighton and Hove Safer Communities Team | Local Authority |  |  |  | 1 | 1 |
| Bury Council | Local Authority |  | 1 |  | 1 | 2 |
| Calderdale Council Highways Department | Local Authority |  |  |  | 1 | 1 |
| Cambridgeshire County Council | Local Authority |  | 1 |  |  | 1 |
| Camden Council | Local Authority |  | 1 |  |  | 1 |
| Canal Trust Bath | Local Authority |  |  |  | 1 | 1 |
| Coventry City Council | Local Authority |  |  |  | 1 | 1 |
| Denbighshire County Council | Local Authority |  |  |  | 1 | 1 |
| Derbyshire County Council | Local Authority |  |  |  | 1 | 1 |
| Dorset Council | Local Authority |  |  |  | 1 | 1 |
| East Midlands Local Education and Training Board | Local Authority |  |  |  | 1 | 1 |
| Hampshire County Council | Local Authority |  | 1 |  |  | 1 |
| Hertfordshire Trading Standards | Local Authority |  | 1 |  |  | 1 |
| Kent County Council | Local Authority |  |  |  | 1 | 1 |
| Leeds City Council | Local Authority | 1 | 1 | 1 |  | 3 |
| Local Medical Committee | Local Authority |  |  |  | 1 | 1 |
| Mayor of London | Local Authority |  | 1 |  |  | 1 |
| Norfolk County Council | Local Authority |  | 2 |  | 1 | 3 |
| Northampton Borough Council | Local Authority |  | 1 |  |  | 1 |
| Northampton County Council | Local Authority |  |  |  | 1 | 1 |
| Nottinghamshire County Council | Local Authority |  |  |  | 1 | 1 |
| Oldham Metropolitan Borough Council | Local Authority | 1 |  |  |  | 1 |
| Plymouth City Council | Local Authority |  |  |  | 1 | 1 |
| Rochdale Metropolitan Borough Council | Local Authority |  |  |  | 1 | 1 |
| Sandwell Borough Council | Local Authority |  |  |  | 1 | 1 |
| Southampton City Council | Local Authority |  |  |  | 1 | 1 |
| Surrey County Council | Local Authority |  |  | 1 |  | 1 |
| Tameside Council | Local Authority |  | 1 |  |  | 1 |
| Telford & Wrekin Council | Local Authority |  | 1 |  |  | 1 |
| Tower Hamlets Council | Local Authority |  |  |  | 1 | 1 |
| ------------------------------------------------------- | Local Service | 1 | 0 | 0 | 8 | 9 |
| Bromley Drug and Alcohol Service | Local Service |  |  |  | 1 | 1 |
| Bury MBC | Local Service |  |  |  | 1 | 1 |
| Drug and Alcohol Action Team Cornwall Council | Local Service |  |  |  | 1 | 1 |
| Forward Trust The Foundary | Local Service |  |  |  | 1 | 1 |
| Local drugs and alcohol team | Local Service |  |  |  | 1 | 1 |
| North Community Mental Health Team | Local Service |  |  |  | 1 | 1 |
| Spectrum Community Health | Local Service |  |  |  | 1 | 1 |
| Transport for London | Local Service | 1 |  |  | 1 | 2 |
| ------------------------------------------------------- | NHS Ambulance | 8 | 14 | 7 | 14 | 43 |
| AACE National Directors of Operations Group | NHS Ambulance |  |  |  | 1 | 1 |
| Association of Ambulance Chief Executives | NHS Ambulance | 1 | 1 | 3 | 3 | 8 |
| East Midlands Ambulance Service | NHS Ambulance |  | 1 |  | 1 | 2 |
| East of England Ambulance Service | NHS Ambulance |  | 1 |  |  | 1 |
| EEAS | NHS Ambulance |  | 1 |  | 1 | 2 |
| London Ambulance Service | NHS Ambulance |  | 1 |  | 1 | 2 |
| National Ambulance Resilience Unit | NHS Ambulance |  | 1 |  |  | 1 |
| North East Ambulance Service | NHS Ambulance |  |  |  | 1 | 1 |
| North West Ambulance Service | NHS Ambulance | 2 | 2 | 1 |  | 5 |
| South Central Ambulance Service NHS Trust | NHS Ambulance | 1 | 1 |  | 2 | 4 |
| South East Coast Ambulance Service | NHS Ambulance | 2 |  |  | 2 | 4 |
| South Western Ambulance Trust | NHS Ambulance |  | 1 | 2 |  | 3 |
| Welsh Ambulance Services NHS Trust | NHS Ambulance | 1 | 4 |  | 2 | 7 |
| West Midlands Ambulance Service | NHS Ambulance | 1 |  |  |  | 1 |
| Yorkshire Ambulance Service | NHS Ambulance |  |  | 1 |  | 1 |
| ------------------------------------------------------- | NHS CCG | 7 | 21 | 4 | 32 | 64 |
| Bath and North East Somerset CCG | NHS CCG |  |  |  | 1 | 1 |
| Birmingham and Solihull CCG | NHS CCG |  | 1 |  | 2 | 3 |
| Blackpool CCG | NHS CCG |  | 1 |  |  | 1 |
| Brighton and Hove CCG | NHS CCG | 1 |  | 2 | 2 | 5 |
| Cambridgeshire and Peterborough CCG | NHS CCG |  |  |  | 3 | 3 |
| CCG Devon | NHS CCG |  |  |  | 1 | 1 |
| Clinical Commissioning Groups | NHS CCG |  | 1 |  |  | 1 |
| East Leicestershire and Rutland CCG | NHS CCG |  | 1 |  |  | 1 |
| Hampshire and Isle of Wight CCG | NHS CCG |  |  |  | 2 | 2 |
| Horsham and Mid Sussex CCG | NHS CCG |  |  |  | 1 | 1 |
| Lincolnshire East CCG | NHS CCG |  |  |  | 1 | 1 |
| Manchester North, Central and South CCGs | NHS CCG |  |  |  | 1 | 1 |
| Mid Essex CCG Trust | NHS CCG | 1 |  |  | 1 | 2 |
| Milton Keynes CCG | NHS CCG |  |  | 1 |  | 1 |
| NHS East Leicestershire and Rutland CGC | NHS CCG |  |  |  | 1 | 1 |
| NHS Hillingdon CCG | NHS CCG |  | 1 |  |  | 1 |
| NHS Kernow CCG | NHS CCG | 1 | 2 |  | 1 | 4 |
| NHS Norfolk and Waveney CCG | NHS CCG |  |  |  | 1 | 1 |
| NHS Northern Eastern and Western Devon CCG | NHS CCG | 1 | 3 |  |  | 4 |
| NHS Oldham CCG | NHS CCG |  | 2 |  | 1 | 3 |
| NHS Sheffield CCG | NHS CCG |  |  |  | 1 | 1 |
| NHS Trafford CCG | NHS CCG |  | 1 |  |  | 1 |
| Nottingham CCG | NHS CCG |  |  |  | 1 | 1 |
| Redbridge CCG | NHS CCG |  |  |  | 1 | 1 |
| Rochdale, Heywood and Middleton CCG | NHS CCG |  | 1 |  |  | 1 |
| Solihull CCG | NHS CCG |  | 1 |  |  | 1 |
| South Tees CCG | NHS CCG |  | 1 |  |  | 1 |
| Stockport CCG | NHS CCG | 1 | 1 |  | 4 | 6 |
| Tameside and Glossop CCG | NHS CCG |  | 2 |  | 3 | 5 |
| Telford and Wrekin CCG | NHS CCG | 1 |  |  |  | 1 |
| Tower Hamlets CCG | NHS CCG |  |  |  | 1 | 1 |
| Trafford CCG | NHS CCG |  | 1 | 1 | 2 | 4 |
| West Norfolk CCG | NHS CCG |  | 1 |  |  | 1 |
| Wigan Borough CCG | NHS CCG | 1 |  |  |  | 1 |
| ------------------------------------------------------- | NHS Community Organization | 0 | 0 | 0 | 4 | 4 |
| ADAPT | NHS Community Organization |  |  |  | 1 | 1 |
| AMHP | NHS Community Organization |  |  |  | 1 | 1 |
| Bristol Community Health | NHS Community Organization |  |  |  | 1 | 1 |
| Derbyshire Community Health Services | NHS Community Organization |  |  |  | 1 | 1 |
| ------------------------------------------------------- | NHS Hospital | 7 | 7 | 3 | 20 | 37 |
| Birmingham Children’s Hospital | NHS Hospital |  | 1 |  |  | 1 |
| Bradford Royal Infirmary | NHS Hospital |  |  |  | 1 | 1 |
| Derriford Hospital | NHS Hospital | 1 |  |  | 1 | 2 |
| Doncaster Royal Infirmary | NHS Hospital | 1 | 1 |  |  | 2 |
| Edgware Community Hospital | NHS Hospital |  |  |  | 1 | 1 |
| Homerton University Hospital | NHS Hospital |  |  |  | 1 | 1 |
| James Paget University Hospital | NHS Hospital | 1 |  |  |  | 1 |
| Luton and Dunstable Hospital | NHS Hospital |  |  |  | 1 | 1 |
| Neville Hall Hospital | NHS Hospital |  |  |  | 1 | 1 |
| Newmarket Community Hospital | NHS Hospital |  |  |  | 1 | 1 |
| Norfolk and Norwich University Hospital | NHS Hospital |  |  | 1 | 1 | 2 |
| North Middlesex Hospital | NHS Hospital |  |  |  | 1 | 1 |
| North Tyneside General Hospital | NHS Hospital |  | 1 |  |  | 1 |
| Northern General Hospital | NHS Hospital |  |  | 1 |  | 1 |
| Pinderfields General Hospital | NHS Hospital |  |  | 1 |  | 1 |
| Princess Alexandra Hospital | NHS Hospital |  |  |  | 1 | 1 |
| Queen Elizabeth Hospital | NHS Hospital |  | 1 |  |  | 1 |
| Royal Bolton Hospital | NHS Hospital |  | 1 |  |  | 1 |
| Royal Free Hospital | NHS Hospital | 1 |  |  |  | 1 |
| Royal Gwent Hospital | NHS Hospital |  | 1 |  |  | 1 |
| Royal Hampshire County Hospital | NHS Hospital |  |  |  | 1 | 1 |
| Royal Stoke University Hospital | NHS Hospital | 1 |  |  | 1 | 2 |
| Southend University Hospital | NHS Hospital |  |  |  | 1 | 1 |
| Springfield Hospital | NHS Hospital |  |  |  | 1 | 1 |
| St Georges Hospital | NHS Hospital | 1 |  |  |  | 1 |
| St Mary’s Hospital | NHS Hospital |  |  |  | 1 | 1 |
| St. Peter's Hospital | NHS Hospital | 1 |  |  |  | 1 |
| Teesside University Hospitals | NHS Hospital |  |  |  | 1 | 1 |
| University Hospital of Wales | NHS Hospital |  |  |  | 1 | 1 |
| West Suffolk Hospital | NHS Hospital |  |  |  | 1 | 1 |
| Weymouth Street Hospital | NHS Hospital |  | 1 |  |  | 1 |
| Wythenshawe Hospital | NHS Hospital |  |  |  | 1 | 1 |
| Yeovil District Hospital | NHS Hospital |  |  |  | 1 | 1 |
| ------------------------------------------------------- | NHS Organization | 9 | 18 | 18 | 50 | 95 |
| NHS Digital | NHS Organization |  | 2 | 1 | 1 | 4 |
| NHS England | NHS Organization | 7 | 12 | 16 | 42 | 77 |
| NHS Improvement | NHS Organization | 1 | 1 | 1 | 4 | 7 |
| NHS Pathways | NHS Organization |  | 1 |  | 1 | 2 |
| NHS Professionals | NHS Organization |  | 1 |  |  | 1 |
| NHS Wales | NHS Organization | 1 | 1 |  | 1 | 3 |
| Primary Care Support England | NHS Organization |  |  |  | 1 | 1 |
| ------------------------------------------------------- | NHS Service | 1 | 0 | 0 | 1 | 2 |
| Blood Transfusion Service | NHS Service | 1 |  |  |  | 1 |
| Clinical Assessment Team | NHS Service |  |  |  | 1 | 1 |
| ------------------------------------------------------- | NHS Trust | 38 | 120 | 20 | 126 | 304 |
| 5 Boroughs Partnership NHS Foundation Trust | NHS Trust | 1 | 2 |  |  | 3 |
| ABMU Health Board | NHS Trust |  |  |  | 1 | 1 |
| Aneurin Bevan University Health Board | NHS Trust |  | 1 |  | 2 | 3 |
| Avon & Wiltshire NHS Mental Health Partnership Trust | NHS Trust | 1 | 1 |  | 1 | 3 |
| Barking, Havering & Redbridge University Hospitals NHS Trust | NHS Trust |  |  | 1 | 1 | 2 |
| Barnsley Hospital NHS Trust | NHS Trust |  | 1 |  |  | 1 |
| Barts Health NHS Foundation Trust | NHS Trust | 2 | 3 | 1 | 3 | 9 |
| BCUHB | NHS Trust |  | 2 | 1 | 2 | 5 |
| Berkshire Healthcare NHS Foundation Trust | NHS Trust | 1 |  |  |  | 1 |
| BHRUT | NHS Trust |  |  |  | 1 | 1 |
| Birmingham & Solihull Mental Health Trust | NHS Trust | 1 | 2 |  | 1 | 4 |
| Birmingham Community Healthcare NHS Trust | NHS Trust |  | 1 |  | 2 | 3 |
| Birmingham Women’s and Children’s NHS Trust | NHS Trust |  | 1 |  | 1 | 2 |
| Black Country NHS Foundation Trust | NHS Trust |  |  |  | 2 | 2 |
| Blackpool Teaching Hospitals NHS Foundation Trust | NHS Trust | 1 | 1 |  | 2 | 4 |
| Bradford District Care Foundation Trust | NHS Trust | 1 |  |  |  | 1 |
| Bradford Teaching Hospitals NHS Trust | NHS Trust |  | 1 |  |  | 1 |
| Brighton & Sussex University Hospitals NHS Trust | NHS Trust | 1 | 1 |  |  | 2 |
| Bristol University Hospitals | NHS Trust |  | 1 |  |  | 1 |
| Cambridge and Peterborough NHS Trust | NHS Trust |  | 1 |  | 2 | 3 |
| Camden & Islington NHS Foundation Trust | NHS Trust |  | 2 | 1 | 2 | 5 |
| Cardiff & Vale University Health Board | NHS Trust |  | 2 | 1 | 1 | 4 |
| Central & North West London NHS Foundation Trust | NHS Trust |  | 3 |  | 2 | 5 |
| Central Manchester University Hospitals NHS Foundation Trust | NHS Trust |  |  |  | 5 | 5 |
| Cornwall Partnership NHS Foundation Trust | NHS Trust |  | 1 | 1 | 1 | 3 |
| Coventry & Warwickshire Partnership NHS Trust | NHS Trust |  | 2 |  | 3 | 5 |
| Cumbria, Northumberland, Tyne & Wear NHS Trust | NHS Trust |  | 1 |  | 1 | 2 |
| Cwm Taf Morgannwg University Health Board | NHS Trust | 1 | 2 |  | 2 | 5 |
| Derby and Burton University Hospitals | NHS Trust |  | 1 |  | 1 | 2 |
| Derbyshire Healthcare NHS Foundation Trust | NHS Trust |  | 1 |  | 1 | 2 |
| Devon Partnership NHS Trust | NHS Trust |  | 3 | 1 | 3 | 7 |
| Dorset Healthcare University NHS Foundation Trust | NHS Trust |  | 1 |  | 1 | 2 |
| Dudley and Walsall Mental Health Partnership NHS Trust | NHS Trust | 1 |  |  |  | 1 |
| Dudley Group NHS Foundation Trust | NHS Trust |  |  | 1 |  | 1 |
| East & North Hertfordshire NHS Trust | NHS Trust | 1 |  |  |  | 1 |
| East Kent Hospitals University NHS Foundation Trust | NHS Trust | 1 | 1 |  | 1 | 3 |
| East Lancashire Healthcare NHS Trust | NHS Trust |  |  |  | 1 | 1 |
| East London NHS Foundation Trust | NHS Trust |  | 4 |  | 1 | 5 |
| East Suffolk and North Essex NHS Foundation Trust | NHS Trust |  | 1 |  |  | 1 |
| Epsom & St Helier University Hospitals NHS Trust | NHS Trust |  | 1 | 1 |  | 2 |
| EPUT | NHS Trust | 1 |  |  | 1 | 2 |
| Frimley Park Hospital NHS Trust | NHS Trust |  | 2 | 1 | 1 | 4 |
| Gloucestershire Care Services NHS Trust | NHS Trust |  |  |  | 1 | 1 |
| Gloucestershire Hospitals NHS Trust | NHS Trust |  | 2 |  |  | 2 |
| Great Western Hospitals NHS Foundation Trust | NHS Trust | 1 | 1 |  | 1 | 3 |
| Greater Manchester Health & Social Care Partnership | NHS Trust | 1 | 3 |  | 4 | 8 |
| Greater Manchester Mental Health NHS Foundation Trust | NHS Trust |  | 4 | 1 | 2 | 7 |
| Guys and St Thomas NHS Trust | NHS Trust | 1 |  |  | 1 | 2 |
| Heart of England NHS Foundation Trust | NHS Trust | 1 | 1 |  | 1 | 3 |
| Hillingdon Hospital NHS Trust | NHS Trust |  |  |  | 1 | 1 |
| Hull and East Yorkshire NHS Trust | NHS Trust |  |  |  | 1 | 1 |
| Isle of Wight NHS Trust | NHS Trust |  | 1 |  | 2 | 3 |
| Kent and Medway NHS and Social Care Partnership Trust | NHS Trust | 2 |  |  | 2 | 4 |
| Kent Community NHS Trust | NHS Trust |  |  |  | 1 | 1 |
| Kings College Hospital NHS Foundation Trust | NHS Trust | 1 |  |  | 2 | 3 |
| Leeds Community Healthcare NHS Trust | NHS Trust |  |  |  | 1 | 1 |
| Leicester University Hospitals NHS Trust | NHS Trust | 1 |  | 1 | 1 | 3 |
| Leicestershire Partnership NHS Trust | NHS Trust |  | 5 |  |  | 5 |
| Lewisham & Greenwich University Hospital NHS Trust | NHS Trust |  | 1 |  |  | 1 |
| Maidstone & Tunbridge Wells NHS Foundation Trust | NHS Trust |  |  | 1 |  | 1 |
| Manchester Health and Social Care Partnership | NHS Trust |  | 1 |  | 1 | 2 |
| Manchester Mental Health NHS Trust | NHS Trust |  |  |  | 1 | 1 |
| Manchester University NHS Foundation Trust | NHS Trust | 1 | 2 |  |  | 3 |
| Medway NHS Foundation Trust | NHS Trust | 1 | 1 |  |  | 2 |
| Mersey Care NHS Trust | NHS Trust |  |  | 1 |  | 1 |
| Mid Yorkshire NHS Trust | NHS Trust |  |  |  | 1 | 1 |
| Midlands Partnership NHS Foundation Trust | NHS Trust |  | 1 |  | 4 | 5 |
| NHS Lothian | NHS Trust |  |  |  | 1 | 1 |
| Norfolk & Suffolk NHS Trust | NHS Trust |  | 2 |  |  | 2 |
| North Cumbria University Hospitals NHS Trust | NHS Trust |  |  | 1 |  | 1 |
| North East London NHS Foundation Trust | NHS Trust | 1 | 2 |  | 2 | 5 |
| North Essex Partnership University NHS Foundation Trust | NHS Trust |  |  |  | 3 | 3 |
| North Midlands NHS Trust | NHS Trust |  |  |  | 1 | 1 |
| North West Anglia NHS Trust | NHS Trust |  | 1 |  | 1 | 2 |
| Northern Care Alliance NHS Group | NHS Trust | 1 | 1 |  | 1 | 3 |
| Northern Health and Social Care Trust | NHS Trust |  |  | 1 |  | 1 |
| Nottingham University Hospitals NHS Trust | NHS Trust |  | 1 |  | 1 | 2 |
| Nottinghamshire Healthcare NHS Foundation Trust | NHS Trust | 1 |  |  | 5 | 6 |
| Oxford Health NHS Foundation Trust | NHS Trust | 2 |  |  |  | 2 |
| Oxleas Mental Health Trust | NHS Trust |  |  |  | 1 | 1 |
| Oxleas NHS Foundation Trust | NHS Trust | 1 | 1 |  | 1 | 3 |
| Pennine Care Mental Health Trust | NHS Trust |  |  |  | 1 | 1 |
| Pennine Care NHS Foundation Trust | NHS Trust | 1 | 9 |  | 5 | 15 |
| Plymouth Hospitals NHS Trust | NHS Trust |  | 1 |  |  | 1 |
| Rotherham Hospital NHS Foundation Trust | NHS Trust |  | 1 |  |  | 1 |
| Royal Bolton Hospital NHS Foundation Trust | NHS Trust |  | 2 |  | 1 | 3 |
| Royal Devon and Exeter NHS Foundation Trust | NHS Trust | 1 |  |  | 1 | 2 |
| Royal Orthopaedic Hospital NHS Foundation Trust | NHS Trust |  | 1 |  |  | 1 |
| Royal Surrey County Hospital NHS Foundation Trust | NHS Trust |  |  |  | 2 | 2 |
| Royal United Hospitals Bath NHS Trust | NHS Trust |  | 1 |  | 1 | 2 |
| Salford Royal Hospital NHS Trust | NHS Trust |  | 1 |  | 2 | 3 |
| Salisbury Hospital NHS Trust | NHS Trust |  | 1 |  |  | 1 |
| Sandwell and West Birmingham NHS Trust | NHS Trust |  | 2 |  |  | 2 |
| Sherwood Forest Hospitals NHS Foundation Trust | NHS Trust |  |  | 1 |  | 1 |
| Shropshire Community Health NHS Trust | NHS Trust | 1 |  |  |  | 1 |
| Solent NHS Trust | NHS Trust |  | 1 |  |  | 1 |
| South Essex Partnership University NHS Foundation Trust | NHS Trust | 1 | 1 |  | 1 | 3 |
| South London and Maudsley NHS Foundation Trust | NHS Trust |  | 1 |  | 4 | 5 |
| South Manchester University Hospital NHS Foundation Trust | NHS Trust | 1 |  |  |  | 1 |
| South Staffordshire & Shropshire Health Foundation Trust | NHS Trust |  |  |  | 2 | 2 |
| South Tees NHS Trust | NHS Trust |  | 1 |  |  | 1 |
| South West Yorkshire Partnership NHS Foundation Trust | NHS Trust |  |  |  | 1 | 1 |
| Southampton University Hospital NHS Foundation Trust | NHS Trust |  |  | 1 |  | 1 |
| Southern Health NHS Foundation Trust of Tatchbury Mount | NHS Trust |  |  |  | 1 | 1 |
| St George’s Mental NHS Trust | NHS Trust |  | 1 |  |  | 1 |
| St Marien Hospital Trust | NHS Trust |  |  |  | 1 | 1 |
| St Peters and Ashford hospitals Chertsey | NHS Trust |  |  |  | 1 | 1 |
| Stockport NHS Foundation Trust | NHS Trust |  | 1 |  |  | 1 |
| Surrey and Borders Partnership NHS Foundation Trust | NHS Trust |  | 2 | 1 |  | 3 |
| Sussex Partnership NHS Foundation Trust | NHS Trust |  | 1 |  | 2 | 3 |
| Tameside & Glossop Integrated Care NHS Foundation Trust | NHS Trust | 1 | 2 |  | 1 | 4 |
| Tees, Esk and Wear Valleys NHS Foundation Trust | NHS Trust | 1 |  |  | 1 | 2 |
| Torbay and South Devon NHS Trust | NHS Trust |  | 1 |  |  | 1 |
| United Lincolnshire Hospitals NHS Trust | NHS Trust | 1 | 1 |  |  | 2 |
| University College London Hospitals NHS Foundation Trust | NHS Trust |  | 1 |  |  | 1 |
| University Hospital Birmingham NHS Foundation Trust | NHS Trust |  | 3 |  | 1 | 4 |
| Walsall and Dudley Mental Health NHS Trust | NHS Trust |  |  | 1 |  | 1 |
| Walsall Healthcare NHS Trust | NHS Trust |  |  |  | 1 | 1 |
| West London Mental Health NHS Trust | NHS Trust |  |  |  | 1 | 1 |
| Western Sussex NHS Trust | NHS Trust |  | 2 |  |  | 2 |
| Worcestershire Health and Care NHS Trust | NHS Trust |  | 2 |  | 1 | 3 |
| Wrightington Wigan & Leigh NHS Trust | NHS Trust |  | 1 |  |  | 1 |
| Wye Valley NHS Trust | NHS Trust |  | 1 |  |  | 1 |
| ------------------------------------------------------- | Pharmacy | 1 | 2 | 0 | 2 | 5 |
| Adams Pharmacy | Pharmacy |  |  |  | 1 | 1 |
| Boots UK Ltd | Pharmacy |  |  |  | 1 | 1 |
| Haverhill Pharmacy | Pharmacy |  | 1 |  |  | 1 |
| Hindley Health Centre Pharmacy | Pharmacy | 1 |  |  |  | 1 |
| United Pharmacy | Pharmacy |  | 1 |  |  | 1 |
| ------------------------------------------------------- | Police | 5 | 14 | 5 | 25 | 49 |
| ACPO | Police |  |  |  | 1 | 1 |
| Association of Police Officers | Police |  |  |  | 1 | 1 |
| Avon and Somerset Police | Police |  | 1 |  |  | 1 |
| Bedfordshire Police | Police | 1 | 1 |  |  | 2 |
| City of London Police | Police | 1 |  |  |  | 1 |
| Cleveland Police | Police |  | 1 |  |  | 1 |
| Devon & Cornwall Police | Police |  |  |  | 1 | 1 |
| Dorset Police | Police |  | 1 |  | 1 | 2 |
| Essex Police | Police |  |  |  | 2 | 2 |
| Greater Manchester Police | Police | 1 | 4 | 1 | 1 | 7 |
| Gwent Police | Police |  |  |  | 1 | 1 |
| Hampshire Police | Police |  |  |  | 1 | 1 |
| Longton Police Station | Police |  |  |  | 1 | 1 |
| Metropolitan Police Service | Police | 1 | 2 | 2 | 4 | 9 |
| Northumbria Police Service | Police |  |  |  | 1 | 1 |
| South Wales Police | Police |  |  |  | 1 | 1 |
| Staffordshire Police | Police |  | 1 | 1 |  | 2 |
| Surrey Police | Police |  | 1 |  | 4 | 5 |
| Sussex Police | Police |  | 1 |  | 2 | 3 |
| Thames Valley Police | Police | 1 |  |  |  | 1 |
| Warwickshire Police | Police |  | 1 |  |  | 1 |
| West Mercia Constabulary | Police |  |  | 1 |  | 1 |
| West Midlands Police | Police |  |  |  | 3 | 3 |
| ------------------------------------------------------- | Prison | 3 | 2 | 4 | 15 | 24 |
| Birmingham Prison | Prison |  |  |  | 1 | 1 |
| Chief Inspector of Prisons | Prison | 1 |  |  |  | 1 |
| Director General of Prisons | Prison |  |  |  | 1 | 1 |
| Governor HMP Stoke Heath | Prison |  |  | 1 |  | 1 |
| HMP Belmarsh and Healthcare UK | Prison |  |  |  | 1 | 1 |
| HMP Berwyn | Prison | 1 |  |  |  | 1 |
| HMP Dovegate | Prison |  | 1 |  |  | 1 |
| HMP Durham | Prison | 1 |  |  | 1 | 2 |
| HMP Exeter | Prison |  |  |  | 1 | 1 |
| HMP Guys Marsh | Prison |  |  |  | 2 | 2 |
| HMP Hewell | Prison |  |  |  | 2 | 2 |
| HMP High Down | Prison |  |  |  | 1 | 1 |
| HMP Humber | Prison |  |  |  | 1 | 1 |
| HMP Liverpool | Prison |  |  |  | 1 | 1 |
| HMP Long Lartin | Prison |  |  | 1 |  | 1 |
| HMP Rochester | Prison |  |  | 1 |  | 1 |
| HMP Winchester | Prison |  |  |  | 1 | 1 |
| HMP Woodhill | Prison |  |  | 1 |  | 1 |
| HMP Wormwood Scrubs | Prison |  | 1 |  | 1 | 2 |
| HMP Wymott | Prison |  |  |  | 1 | 1 |
| ------------------------------------------------------- | Professional Body | 24 | 29 | 15 | 91 | 159 |
| BMA | Professional Body |  |  |  | 3 | 3 |
| BNF | Professional Body |  |  |  | 1 | 1 |
| British Association of Perinatal Medicine | Professional Body |  | 1 |  |  | 1 |
| British Orthopaedic Association | Professional Body |  | 1 |  |  | 1 |
| British Renal Society | Professional Body |  |  |  | 1 | 1 |
| Chief Coroner | Professional Body |  |  |  | 1 | 1 |
| Civil Aviation Authority | Professional Body |  | 1 |  |  | 1 |
| College of Emergency Medicine | Professional Body |  |  |  | 1 | 1 |
| College of Policing | Professional Body | 2 | 4 |  | 4 | 10 |
| Company Chemists’ Association. | Professional Body | 1 |  |  |  | 1 |
| CQC | Professional Body | 3 | 3 | 3 | 11 | 20 |
| Devon Local Medical Committee | Professional Body |  |  | 1 | 2 | 3 |
| Difficult Airway Society | Professional Body |  | 1 |  |  | 1 |
| Dispensing Doctors Association | Professional Body |  |  |  | 1 | 1 |
| Emergency Call Prioritization Advisory Group | Professional Body |  |  |  | 2 | 2 |
| Faculty of Intensive care of Royal College of Anaesthetists | Professional Body |  |  |  | 1 | 1 |
| Fire Officers Association | Professional Body |  |  |  | 1 | 1 |
| General Dental Council | Professional Body |  |  |  | 1 | 1 |
| General Medical Council | Professional Body | 2 | 2 | 1 | 4 | 9 |
| General Pharmaceutical Council | Professional Body | 1 | 3 |  | 3 | 7 |
| Intensive care society | Professional Body |  |  |  | 1 | 1 |
| Joint Royal Colleges Ambulance Liaison Committee | Professional Body | 1 |  |  | 2 | 3 |
| MHRA | Professional Body | 3 | 3 | 5 | 15 | 26 |
| National Association of Ambulance Medical Directors | Professional Body | 1 |  |  | 2 | 3 |
| National Patient Safety Agency | Professional Body |  |  |  | 3 | 3 |
| National Police Chiefs' Council | Professional Body | 1 | 2 |  | 7 | 10 |
| National Probation Service | Professional Body |  | 1 |  | 1 | 2 |
| NICE | Professional Body | 4 |  |  | 9 | 13 |
| Nursing and Midwifery Council | Professional Body | 1 |  | 1 |  | 2 |
| Renal Association | Professional Body |  |  |  | 1 | 1 |
| Resuscitation Council UK | Professional Body | 1 |  |  |  | 1 |
| Royal College of Anaesthetists | Professional Body |  | 1 |  | 1 | 2 |
| Royal College of Emergency Medicine | Professional Body |  |  |  | 1 | 1 |
| Royal College of General Practitioners | Professional Body |  | 2 |  | 1 | 3 |
| Royal College of Nursing | Professional Body |  |  |  | 1 | 1 |
| Royal College of Obstetrics & Gynaecology | Professional Body |  |  | 1 |  | 1 |
| Royal College of Paramedics | Professional Body |  |  |  | 1 | 1 |
| Royal College of Pathologists | Professional Body |  |  |  | 2 | 2 |
| Royal College of Physicians | Professional Body |  |  | 1 | 1 | 2 |
| Royal College of Psychiatrists | Professional Body | 1 |  |  | 1 | 2 |
| Royal College of Surgeons | Professional Body |  | 2 |  | 1 | 3 |
| Royal Pharmaceutical Society | Professional Body | 2 | 1 | 1 | 2 | 6 |
| TOXBASE | Professional Body |  | 1 |  |  | 1 |
| UK Homecare Association | Professional Body |  |  | 1 |  | 1 |
| Vascular Access Society of Britain & Ireland | Professional Body |  |  |  | 1 | 1 |
| ------------------------------------------------------- | Professional Individual | 1 | 0 | 0 | 5 | 6 |
| Director of Public Prosecutions | Professional Individual |  |  |  | 1 | 1 |
| Justice Regional Team Chair | Professional Individual |  |  |  | 1 | 1 |
| Landlord | Professional Individual | 1 |  |  |  | 1 |
| Policy and Patient Safety Directorate | Professional Individual |  |  |  | 1 | 1 |
| Senior Coroner Sunderland | Professional Individual |  |  |  | 1 | 1 |
| Senior Pastor and Head of Safeguarding | Professional Individual |  |  |  | 1 | 1 |
| ------------------------------------------------------- | Social Charity | 2 | 2 | 0 | 6 | 10 |
| Change Grow Live | Social Charity |  |  |  | 2 | 2 |
| Phoenix Futures | Social Charity |  |  |  | 1 | 1 |
| Skillsforcare | Social Charity |  |  |  | 1 | 1 |
| Stoneham Bass | Social Charity |  | 1 |  |  | 1 |
| Trading Standards Institute | Social Charity | 1 |  |  |  | 1 |
| Turning Point | Social Charity |  | 1 |  | 1 | 2 |
| Victory Outreach Manchester | Social Charity |  |  |  | 1 | 1 |
| Wallich Centre | Social Charity | 1 |  |  |  | 1 |
| ------------------------------------------------------- | Unclassified | 0 | 0 | 0 | 7 | 7 |
| [REDACTED] | Unclassified |  |  |  | 7 | 7 |

***References***

1.	Aronson JK, Ferner RE. Clarification of terminology in drug safety [Internet]. Vol. 28, Drug Safety. Drug Saf; 2005 [cited 2021 Jun 22]. p. 851–70. Available from: https://pubmed.ncbi.nlm.nih.gov/16180936/

2.	Misuse of Drugs Act 1971. Statute Law Database; 1971.

3.	Whitting E. Prevention of Future Deaths Report 2020-0144 [Internet]. 2020 [cited 2021 Jun 28]. Available from: https://www.judiciary.uk/publications/jerrelle-mckenzie/

4.	Agriculture (Poisonous Substances) Act 1952. 1952.
